# Paediatric infection hospital admissions in England before, during and after the COVID-19 pandemic

**DOI:** 10.1101/2025.11.04.25339490

**Authors:** Charlotte Jackson, Linda Wijlaars, Fariyo Abdullahi, Mengyun Liu, Ali Judd, Claire Thorne, Intira Jeannie Collins, Costanza Di Chiara, Pia Hardelid

**Affiliations:** MRC Clinical Trials Unit at UCL, 90 High Holborn, London WC1V 6LJ, UK; UCL Great Ormond Street Institute for Child Health, 30 Guilford Street, London WC1N 1EH, UK; Fondazione Penta ETS, Padua, Italy; Department for Women’s and Children’s Health, University of Padova, Via Giustiniani, 3 - 35128 Padua, Italy; National Institute for Health Research Great Ormond Street Hospital Biomedical Research Centre

## Abstract

The COVID-19 pandemic and associated non-pharmaceutical interventions (NPIs: containment measures/”lockdown”) reduced transmission of other infections. We quantified changes in hospital admission rates for respiratory and gastrointestinal infections amongst young children in England during and after implementation of NPIs, compared to pre-pandemic, and variations by sociodemographic and clinical characteristics. Children aged <5 years at any time between 1 January 2017 and 31 January 2022 were followed from birth or 1 January 2017, until their 5^th^ birthday, death or 31 January 2022, in a birth cohort based on Hospital Episode Statistics data. Quarterly emergency admission rates for respiratory and gastrointestinal infections from April-June 2020 onwards were compared to corresponding quarters in 2017-19 using Poisson regression models, with and without interaction terms for time period and sociodemographic/clinical characteristics. Admission rates for respiratory and gastrointestinal infections were lower in April-June 2020 compared to this quarter pre-pandemic (incidence rate ratio (99% CI) 0.17 (0.17-0.18) for respiratory; 0.29 (0.28-0.31) for gastrointestinal). Rates remained below pre-pandemic levels until April-June 2021 (respiratory infections) and July-September 2021 (gastrointestinal infections), subsequently increasing above the corresponding pre-pandemic quarters. Changes in rates did not differ by sociodemographic/clinical characteristics. These results can inform public health planning for future pandemics and their aftermath.

## Introduction

Following the emergence of SARS-CoV-2 in December 2019 and the ensuing COVID-19 pandemic, non-pharmaceutical interventions (NPIs) such as closure of schools, nurseries, shops and restaurants, and directives and advice regarding working from home, restricted socializing and mask wearing, were widely used to slow viral spread. These measures reduced SARS-CoV-2 transmission [1] and were followed by reductions in the incidence of other respiratory infections in diverse settings [2–4], with subsequent resurgences after NPIs were relaxed [5]. Although changes in testing may partially explain changes in reported incidence of infections [6], active surveillance in households also found lower incidence of respiratory virus infections during the pandemic [7]. Laboratory isolations of gastrointestinal (GI) viruses, particularly norovirus, also declined while NPIs were active [8, 9].

In England, NPIs were first implemented in late March 2020 [10]. Substantial reductions in the number of hospital admissions amongst children aged <15 years in England have been reported for multiple infections in 2020-21 compared to 2017-20, with some evidence of resurgence up to May 2021 following the removal of NPIs [11]. Declines occurred in all regions, and deprivation, ethnic and comorbidity groups [11].

Besides disrupting pathogen transmission, the COVID-19 pandemic and associated NPIs also affected parents’ propensity to seek healthcare for unwell children [12], hospital admission thresholds, and childhood vaccination programmes and uptake [13]. These factors will contribute to changes in hospital admission rates, which may differ with sociodemographic and clinical characteristics [14]. Understanding these impacts will inform planning for changes in healthcare demand during and after any future pandemic.

In this study, we compare hospital admission rates for respiratory and GI infections before and during the pandemic amongst young children in England. We add to previous evidence by focusing on the most vulnerable age group of <5 years, with a longer study period, and assess whether impacts on admissions in this age group differed according to children’s social and demographic characteristics.

## Methods

This analysis utilizes a birth cohort based on Hospital Episode Statistics (HES) birth admission data [15], linked to HES Admitted Patient Care (APC) data [16], to estimate rates of infection admissions amongst children aged <5 years. HES is an administrative database covering all hospital admissions in National Health Service (NHS) hospitals in England.

### Data sources

The birth cohort was assembled from HES data, comprising all singleton children born alive in NHS hospitals in England during 1998-2022 (∼97% of births in England) [15]. The cohort dataset is linked to Office for National Statistics birth registrations and mortality data, and NHS birth notification data [17]. The birth cohort was linked to an extract of HES APC data containing admissions for infections/infectious syndromes from January 2017 to January 2022. HES APC contains data on “finished consultant episodes” (FCEs) each representing a period of care under one consultant; an admission may comprise multiple FCEs [16].

### Study population and follow-up

The study period was 1 January 2017 to 31 January 2022. Children were included if they were aged <5 years on 1 January 2017 or born after that date, and were followed from the later of 1 January 2017 or their birthdate until the earliest of their 5^th^ birthday, death or 31 January 2022. Children were considered at risk of hospitalization for the outcomes of interest throughout their follow-up except during hospitalization for these outcomes; all children were included irrespective of their duration of follow-up.

### Outcomes

Outcomes of interest were emergency hospital admissions with any International Classification of Disease 10 (ICD-10) diagnostic code reflecting a) respiratory or b) GI infection as the primary diagnosis of the first FCE of the admission. An admission was defined as a continuous stay in hospital consisting of one or more FCEs. Re-admissions within one day were counted as a single admission. Children could contribute multiple admissions to the analysis.

We used published code lists to identify relevant ICD-10 codes [18] (Appendix Table 1). Emergency admissions were identified from the admission method as defined in the HES data dictionary [19]. Attendances at Accident & Emergency (A&E) without subsequent admission are not included in HES APC. We also assessed emergency admission rates for the specific diagnostic codes accounting for the greatest numbers of FCEs amongst children aged <5 years within the respiratory and GI lists (based on three-character ICD-10 codes recorded as the primary diagnosis), as recorded in HES data for 2018-19 [20] (five codes for respiratory and four for GI infections due to smaller numbers, see Table 1).

**Table 1:** Most common ICD-10 codes indicating respiratory and gastrointestinal infections.

| Group | Code | Diagnosis |
| --- | --- | --- |
| Respiratory<br>infection | J03 | Acute tonsillitis |
|  | J05 | Acute obstructive laryngitis [croup] and epiglottitis |
|  | J06 | Acute upper respiratory infections of multiple and unspecified sites |
|  | J21 | Acute bronchiolitis |
|  | J22 | Unspecified acute lower respiratory infection |
| Gastrointestinal<br>infection | A02 | Other salmonella infections |
|  | A08 | Viral and other specified intestinal infections |
|  | A09 | Other gastroenteritis and colitis of infectious and unspecified origin |
|  | R11 | Nausea and vomiting |

### Exposure variables

Our main exposure of interest was calendar time, defined as before or after the onset of the pandemic / implementation of NPIs (taken as April 2020) and further stratified by quarter (see below).

Sociodemographic and clinical exposures of interest were sex, age, ethnicity, area-level deprivation, preterm birth (<37 weeks) and presence of congenital anomalies. Age was categorized as <3 months, 3-<6 months, 6-<12 months, and 1-<5 years. Ethnicity recorded at birth was categorized into five broad groups used in the UK 2021 Census: Asian or Asian British; Black, Black British, Caribbean or African; Mixed or multiple ethnic groups; White; Any other ethnic group [21]. Area-level deprivation was measured using quintiles of the index of multiple deprivation (IMD) 2010 as supplied in HES. IMD combines indicators of deprivation across seven domains (Income Deprivation, Employment Deprivation, Health Deprivation and Disability, Education Skills and Training Deprivation, Barriers to Housing and Services, Living Environment Deprivation, and Crime) and is measured at the level of Lower layer Super Output Area, a geographic area with approximately 1500 people [22]. IMD was based on the child’s mother’s address at the time of delivery. Children with congenital anomalies were identified as those with a diagnosis recorded in HES, or cause of death before age 1 year recorded in HES or ONS data, of any congenital anomaly expected to result in a child needing medical follow-up for >12 months in ≥50% of cases [23].

### Statistical analysis

Characteristics of children with and without a respiratory (or, separately, GI) infection admission during the study period were summarized. Monthly infection admission rates (all respiratory, all GI, and specific codes) were calculated with 99% confidence intervals (CIs), overall and stratified by each sociodemographic and clinical variable. For graphical presentation, months were classified as falling within periods of national “lockdown” (April 2020 to March 2021) or less restrictive NPIs (July 2020 to June 2021, including a less stringent lockdown in autumn 2020 during which schools remained open and socialising was slightly less restricted than at the start of the first lockdown) [10].

To quantify changes in admission rates over time, we conducted analyses using aggregated quarterly rates. We used Poisson regression models to estimate incidence rate ratios (IRRs) comparing rates in each pandemic quarter to the overall rate across the corresponding pre-pandemic quarters (e.g. rate in April-June 2020 versus rate in April-June in 2017, 2018 and 2019). An IRR <1.0 indicates that admission rates were lower during the given pandemic quarter than in the corresponding pre-pandemic quarter (i.e. lower than expected for the time of year). Additionally, for overall respiratory and GI infection admissions, we fitted models with effect modification terms between the exposure (pre-versus post-pandemic onset) and each of the sociodemographic and clinical characteristics mentioned above, to estimate stratum-specific IRRs and assess whether these indicated differential impact between groups. These were compared to models without effect modification (including only children with data available on the characteristic of interest) using Akaike’s Information Criterion (AIC) with a difference in AIC of >10 considered strong support for the model with the lower value [24]. In the main analyses, repeat admissions for the same child were treated as independent events.

Finally, we estimated the cumulative incidence of first hospitalization with a respiratory or GI infection, by year of birth, for children followed from birth (born in 2017 or later).

All regression analyses used a complete case approach (i.e. included only children with data on the variables included in the respective model). For variables with >10% missing data, we summarized characteristics of children with and without data. 99% CIs were used due to the large sample size and multiple testing.

All analyses were conducted in Stata, version 18.

### Sensitivity analyses

We repeated the regression analyses of total respiratory or GI infection admissions 1) including all (primary and other) diagnoses, recorded in any episode of an admission, and 2) excluding children born in London (a proxy for living in London), due to differences between London and other regions in admission rates [25] and epidemiology of COVID-19 [26]. Finally, we conducted an individual-level analysis of respiratory infection hospitalizations using Poisson regression with cluster-robust standard errors to account for repeat admissions.

### Ethics approval and consent to participate

This study used NHS Hospital Episode Statistics data which was provided within the terms of a data sharing agreement (DARS-NIC-393510-D6H1D-v9.3) by NHS England.

## Results

A total of 5,984,261 children were included in the analysis, of whom 551,505 (9%) and 101,842 (2%) were admitted to hospital at least once with a primary diagnostic code indicating a respiratory or GI infection, respectively, during follow-up. Median [IQR] duration of follow-up was 31 [15–46] months.

Amongst those admitted, the median number of respiratory admissions was 1 [IQR 1-2] and GI admissions also 1 [IQR 1-1]; 2% of all children in the study population had >1 respiratory and 0.1% >1 GI infection admission. Baseline characteristics of children with and without respiratory and GI admissions are shown in Table 2, and characteristics at time of admission (803,550 respiratory and 112,336 GI admissions) in Table 3. Data were missing for >10% of children for IMD and gestational age. There were no clear differences in clinical or sociodemographic characteristics between those with and without data on gestational age, whilst those without data on IMD appeared less likely to have congenital anomalies, be born preterm, have a mother born outside the UK, and have fewer respiratory admissions and GI admissions (Appendix Tables 2 and 3).

**Table 2:** Characteristics of children with and without at least one respiratory or gastrointestinal admission.

|  | Any admission with respiratory code as primary diagnosis |  | Any admission with GI code as primary diagnosis |  |
| --- | --- | --- | --- | --- |
|  | No<br>5,432,756 | Yes<br>551,505 | No<br>5,882,419 | Yes<br>101,842 |
| Sex (n = 5,982,720) |  |  |  |  |
| Male | 2,748,279 (50.6%) | 321,098 (58.2%) | 3,015,944 (51.3%) | 53,433 (52.5%) |
| Female | 2,683,082 (49.4%) | 230,261 (41.8%) | 2,864,973 (48.7%) | 48,370 (47.5%) |
| Congenital anomalies (n = 5,984,261) |  |  |  |  |
| No | 5,275,101 (97.1%) | 514,815 (93.3%) | 5,697,056 (96.8%) | 92,860 (91.2%) |
| Yes | 157,655 (2.9%) | 36,690 (6.7%) | 185,363 (3.2%) | 8,982 (8.8%) |
| Ethnicity (n = 5,544,132) |  |  |  |  |
| White | 3,730,418 (74.1%) | 391,678 (76.6%) | 4,050,339 (74.3%) | 71,757 (75.9%) |
| Asian or Asian British | 614,351 (12.2%) | 61,169 (12.0%) | 663,797 (12.2%) | 11,723 (12.4%) |
| Black, Black British, Caribbean or African | 255,283 (5.1%) | 19,594 (3.8%) | 270,987 (5.0%) | 3,890 (4.1%) |
| Mixed or multiple ethnic groups | 299,037 (5.9%) | 28,251 (5.5%) | 322,463 (5.9%) | 4,825 (5.1%) |
| Any other ethnic group | 133,490 (2.7%) | 10,861 (2.1%) | 142,055 (2.6%) | 2,296 (2.4%) |
| Preterm (n = 4,696,490) |  |  |  |  |
| Term | 4,001,171 (93.9%) | 390,273 (89.6%) | 4,319,263 (93.6%) | 72,181 (89.6%) |
| Preterm | 259,514 (6.1%) | 45,532 (10.4%) | 296,685 (6.4%) | 8,361 (10.4%) |
| Maternal age at delivery (years) (n = 4,203,342) | 30 [26 - 34] | 29 [25 - 33] | 30 [26 - 34] | 29 [24 - 33] |
| Mother born outside UK (n = 3,495,873) |  |  |  |  |
| UK | 2,113,949 (69.8%) | 362,387 (77.4%) | 2,412,432 (70.7%) | 63,904 (75.5%) |
| Non UK | 913,590 (30.2%) | 105,947 (22.6%) | 998,839 (29.3%) | 20,698 (24.5%) |
| IMD quintile (n = 4,779,452) |  |  |  |  |
| Quintile 1 (most deprived) | 1,168,967 (27.4%) | 147,539 (29.1%) | 1,287,685 (27.5%) | 28,821 (30.6%) |
| Quintile 2 | 957,388 (22.4%) | 112,038 (22.1%) | 1,048,373 (22.4%) | 21,053 (22.4%) |
| Quintile 3 | 804,026 (18.8%) | 94,027 (18.6%) | 880,643 (18.8%) | 17,410 (18.5%) |
| Quintile 4 | 694,201 (16.2%) | 80,797 (15.9%) | 760,619 (16.2%) | 14,379 (15.3%) |
| Quintile 5 (least deprived) | 647,992 (15.2%) | 72,477 (14.3%) | 708,002 (15.1%) | 12,467 (13.2%) |
| Total number of respiratory or GI admissions* (n = 551,505; 101,842) |  |  |  |  |
| 1 |  | 411,453 (74.6%) |  | 93,825 (92.1%) |
| 2 |  | 86,769 (15.7%) |  | 6,546 (6.4%) |
| 3 |  | 28,383 (5.1%) |  | 996 (1.0%) |
| 4 |  | 11,706 (2.1%) |  | 238 (0.2%) |
| ≥5 |  | 13,194 (2.4%) |  | 237 (0.2%) |
| Total number of respiratory or GI admissions* (n = 551,505; 101,842) (median [IQR]) |  | 1 [1 - 2] |  | 1 [1 - 1] |

\* Number of respiratory infection admissions for those admitted with respiratory infections; number of GI admissions for those admitted with GI infections.

**Table 3:**
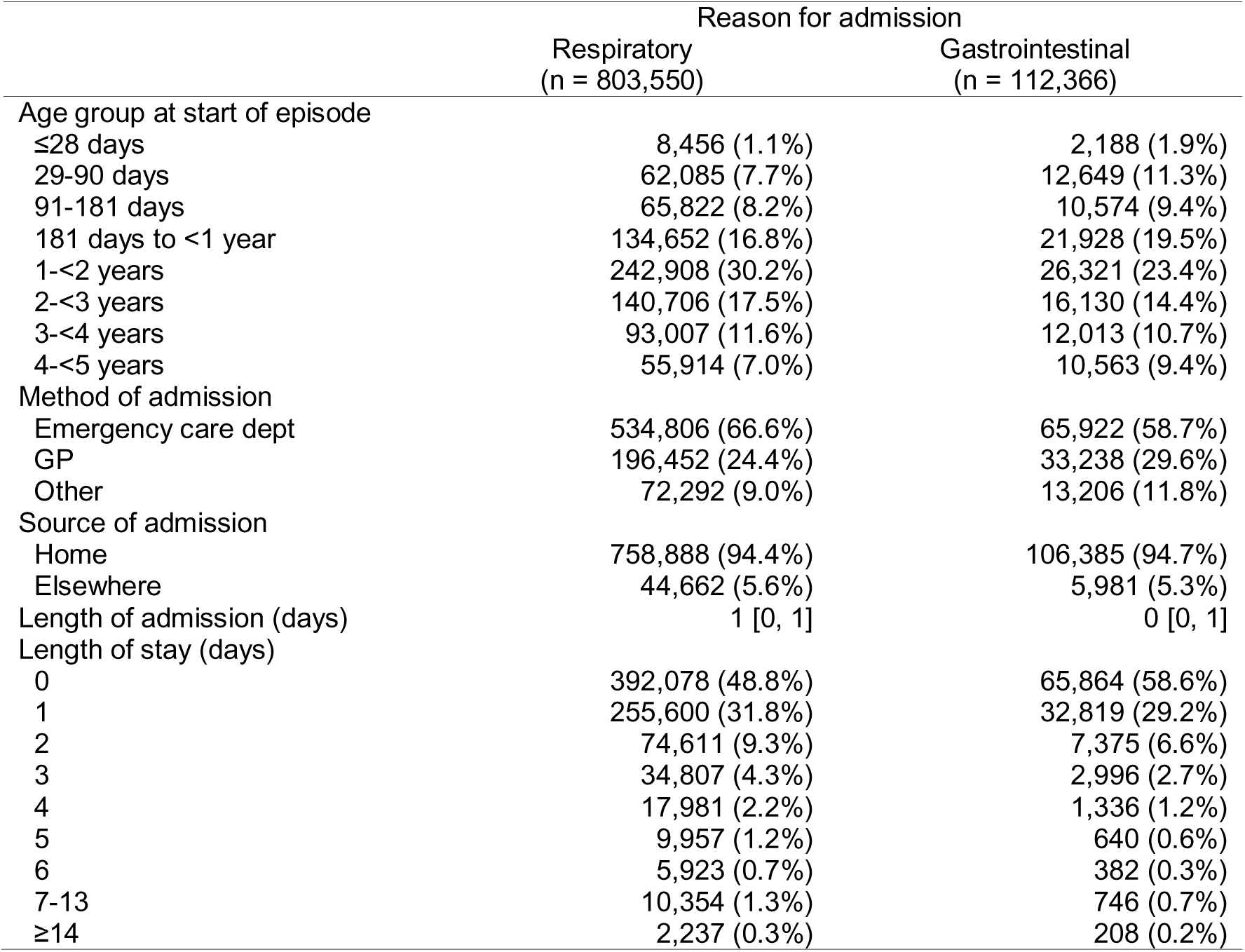
Characteristics of children at time of admission for respiratory and gastrointestinal diagnoses.

### Overall trends in respiratory infection admission rates

Respiratory infection admission rates were declining before the pandemic, fell below the pre-pandemic minimum during the initial lockdown period, and increased through the subsequent period of less restrictive NPIs to a small, off-season peak in September 2020 (Figure 1A). Rates subsequently increased, but with a less clear seasonal pattern than in the pre-pandemic period, during which clear peak in December was evident. Trends for specific respiratory infection admissions were generally similar (Appendix Figures 1-6), although peaks occurred earlier during the final period of NPIs for acute upper respiratory infections of multiple and unspecified sites (Appendix Figure 2), acute tonsillitis (Appendix Figure 4) and acute obstructive laryngitis (croup) (Appendix Figure 5).

**Figure 1:**
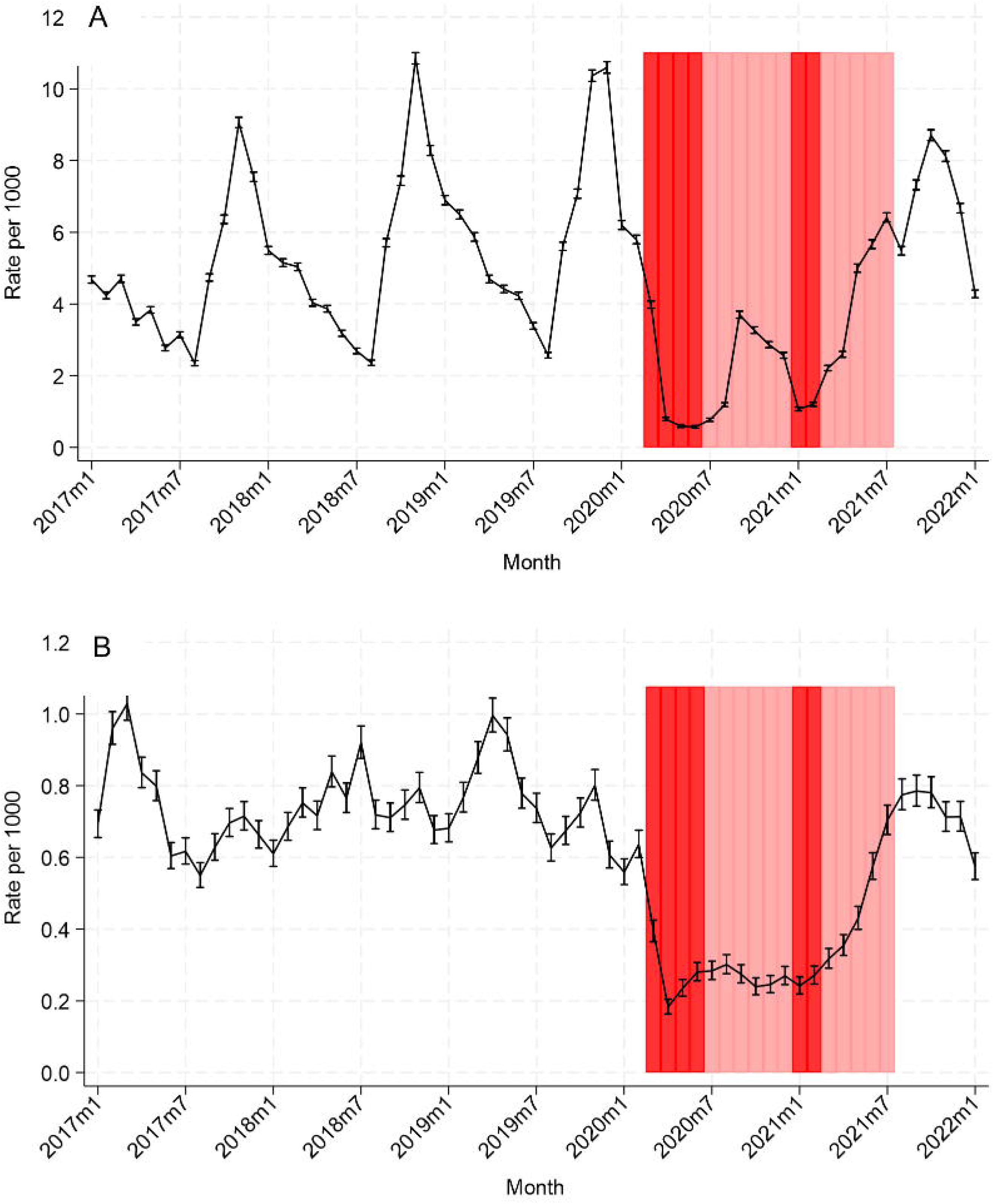
Rates (per 1000 person-months) of hospital admission in children aged <5 years with a primary diagnostic code indicating A) respiratory infection and B) gastrointestinal infection, England, January 2017 to January 2022. Note the differing y axis scales. Red bars show the approximate timing of national lockdowns, pink bars show other NPIs.

**Figure 2:**
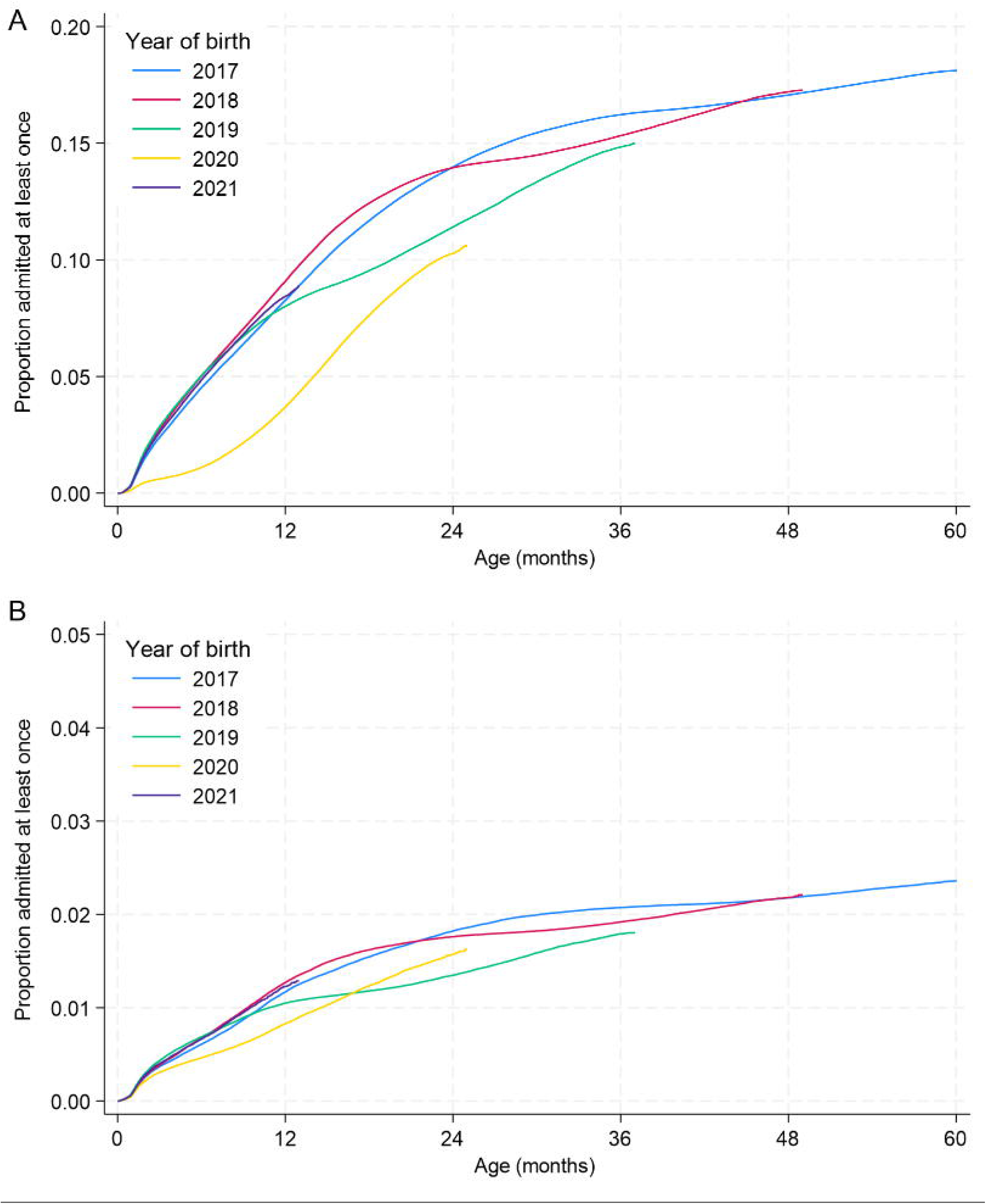
Cumulative incidence, by age, of first hospital admission with a primary diagnostic code indicating A) a respiratory infection or B) a gastrointestinal infection, by year of birth. Note the differing y axis scales.

In April-June 2020, respiratory infection admission rates were reduced by 83% (IRR 0.17, 99% CI 0.17-0.18) compared to April-June 2017-19 (Table 4). Rates remained lower than expected for the time of year until April-June 2021, when rates were higher than in the corresponding pre-pandemic periods (IRR 1.18, 99% CI 1.16-1.20); and increased further by July-September 2021 with IRR 1.77 (99% CI 1.74-1.80). By October-December 2021, rates declined to slightly lower than pre-pandemic levels (IRR 0.90, 99% CI 0.89-0.91).

**Table 4:** Incidence rate ratios comparing quarterly admission rates with primary diagnostic codes indicating respiratory or gastrointestinal infections with corresponding pre-pandemic quarters.

|  | Respiratory infections |  | Gastrointestinal infections |  |
| --- | --- | --- | --- | --- |
|  | IRR | (99% CI) | IRR | (99% CI) |
| Apr-Jun 2020 | 0.17 | (0.17-0.18) | 0.29 | (0.28-0.31) |
| Jul-Sep 2020 | 0.52 | (0.51-0.54) | 0.41 | (0.39-0.44) |
| Oct-Dec 2020 | 0.33 | (0.33-0.34) | 0.35 | (0.33-0.37) |
| Jan-Mar 2021 | 0.29 | (0.28-0.30) | 0.36 | (0.34-0.38) |
| Apr-Jun 2021 | 1.18 | (1.16-1.20) | 0.57 | (0.54-0.59) |
| Jul-Sep 2021 | 1.77 | (1.74-1.80) | 1.11 | (1.07-1.15) |
| Oct-Dec 2021 | 0.90 | (0.89-0.91) | 1.02 | (0.98-1.06) |

Comparing admission rates in April-June 2020 to corresponding pre-pandemic quarters for specific respiratory infections, reductions were greatest for acute bronchiolitis, croup and epiglottitis, and viral wheeze (IRR <0.10 in all cases, Table 5). Changes in IRRs over time for acute bronchiolitis, acute upper respiratory infections of multiple and unspecified sites, and unspecified acute lower respiratory infection generally reflected those seen for all respiratory infection admissions, although the IRR for acute bronchiolitis in the post-NPI period of July-September 2021 was particularly high (5.67, 99% CI 5.47-5.87). Patterns for acute tonsillitis, croup and epiglottitis, and viral wheeze varied. For acute tonsillitis, IRRs were fairly constant between April-June 2020 and January-March 2021, and increased only slightly above 1 in July-September 2021. Croup admission rates were lower than pre-pandemic periods for all quarters except for April-June 2021 (IRR 2.61 (99% CI 2.46-2.77)). Viral wheeze was the only respiratory admission for which rates remained higher in October-December 2021 compared to corresponding pre-pandemic quarters (IRR 1.13, 99% CI 1.10-1.15). All monthly rates since the onset of the pandemic were lower than the pre-pandemic peaks.

**Table 5:** Incident rate ratios comparing quarterly admission rates with specific primary diagnostic codes indicating respiratory infections with corresponding pre-pandemic quarters.

|  | Acute bronchiolitis (J21) |  | Acute upper respiratory infections of multiple and unspecified sites (J06) |  | Unspecified acute lower respiratory infection (J22) |  | Acute tonsillitis (J03) |  | Acute obstructive laryngitis [croup] and epiglottitis (J05) |  | Viral wheeze (B34.9 and R06.2) |  |
| --- | --- | --- | --- | --- | --- | --- | --- | --- | --- | --- | --- | --- |
|  | IRR | 99% CI | IRR | 99% CI | IRR | 99% CI | IRR | 99% CI | IRR | 99% CI | IRR | 99% CI |
| Apr-Jun 2020 | 0.09 | (0.08-0.11) | 0.26 | (0.24-0.28) | 0.15 | (0.13-0.17) | 0.30 | (0.28-0.32) | 0.07 | (0.05-0.09) | 0.04 | (0.03-0.05) |
| Jul-Sep 2020 | 0.56 | (0.52-0.60) | 0.56 | (0.53-0.59) | 0.41 | (0.38-0.45) | 0.43 | (0.41-0.47) | 0.28 | (0.26-0.31) | 0.57 | (0.55-0.59) |
| Oct-Dec 2020 | 0.13 | (0.13-0.14) | 0.31 | (0.30-0.33) | 0.18 | (0.17-0.19) | 0.43 | (0.40-0.45) | 0.14 | (0.12-0.15) | 0.73 | (0.71-0.75) |
| Jan-Mar 2021 | 0.12 | (0.11-0.13) | 0.33 | (0.31-0.35) | 0.15 | (0.14-0.17) | 0.43 | (0.41-0.46) | 0.48 | (0.44-0.52) | 0.31 | (0.30-0.33) |
| Apr-Jun 2021 | 1.03 | (0.98-1.09) | 1.30 | (1.26-1.34) | 1.01 | (0.96-1.06) | 1.02 | (0.97-1.06) | 2.61 | (2.46-2.77) | 0.97 | (0.94-1.00) |
| Jul-Sep 2021 | 5.67 | (5.47-5.87) | 1.37 | (1.32-1.42) | 2.15 | (2.06-2.24) | 1.06 | (1.01-1.11) | 0.41 | (0.38-0.44) | 1.32 | (1.29-1.36) |
| Oct-Dec 2021 | 0.80 | (0.78-0.82) | 0.79 | (0.76-0.81) | 0.93 | (0.90-0.97) | 0.81 | (0.77-0.85) | 0.32 | (0.30-0.35) | 1.13 | (1.10-1.15) |

### Overall trends in GI infection admission rates

GI infection admission rates declined substantially following implementation of NPIs (Figure 1B). The IRR comparing April-June 2020 to April-June 2017-19 was 0.29 (99% CI 0.28-0.31) (Table 4). Rates remained lower than expected until July-September 2021 (IRR 1.11 (99% CI 1.07-1.15)); the following quarter, rates were similar to the corresponding pre-pandemic quarters (IRR 1.02, 99% CI 0.98-1.06).

Specific GI admission rates are shown in Appendix Figures 7-10. The initial lockdown in April-June 2020 had the greatest impact on viral and other specified intestinal infections (IRR 0.17, 99% CI 0.16-0.20, Table 6). Temporal patterns in IRRs were similar for three of the four GI admission codes, but for other salmonella infections IRRs were more varied with wide CIs (Table 6). Admission rates for viral and other specified intestinal infections, other gastroenteritis and colitis of infectious and unspecified origin, and nausea and vomiting remained low up to and including April-June 2021 and increased slightly in the following quarter. In October-December 2021, rates were similar to the corresponding pre-pandemic quarters except for other gastroenteritis and colitis of infectious and unspecified origin (IRR 1.09, 95% CI 1.04-1.14).

**Table 6:** Incidence rate ratios comparing quarterly admission rates with specific primary diagnostic codes indicating gastrointestinal infections with corresponding pre-pandemic quarters.

|  | Viral and other specified<br>intestinal infections (A08) |  | Other gastroenteritis and colitis<br>of infectious and unspecified<br>origin (A09) |  | Other salmonella<br>infections (A02) |  | Nausea and vomiting (R11) |  |
| --- | --- | --- | --- | --- | --- | --- | --- | --- |
|  | IRR | 99% CI | IRR | 99% CI | IRR | 99% CI | IRR | 99% CI |
| Apr-Jun 2020 | 0.17 | (0.16-0.20) | 0.31 | (0.28-0.34) | 0.49 | (0.21-1.14) | 0.52 | (0.46-0.57) |
| Jul-Sep 2020 | 0.25 | (0.22-0.28) | 0.43 | (0.39-0.47) | 1.21 | (0.72-2.02) | 0.66 | (0.60-0.73) |
| Oct-Dec 2020 | 0.21 | (0.19-0.24) | 0.33 | (0.30-0.37) | 0.69 | (0.32-1.48) | 0.62 | (0.56-0.69) |
| Jan-Mar 2021 | 0.18 | (0.16-0.20) | 0.38 | (0.35-0.42) | 1.46 | (0.74-2.88) | 0.71 | (0.64-0.78) |
| Apr-Jun 2021 | 0.43 | (0.39-0.46) | 0.54 | (0.50-0.58) | 1.06 | (0.57-1.97) | 0.87 | (0.80-0.95) |
| Jul-Sep 2021 | 1.06 | (1.00-1.13) | 1.09 | (1.02-1.16) | 1.20 | (0.72-2.02) | 1.18 | (1.09-1.28) |
| Oct-Dec 2021 | 0.98 | (0.92-1.04) | 1.09 | (1.02-1.16) | 0.91 | (0.46-1.81) | 0.94 | (0.86-1.03) |

### Trends in admission rates by sociodemographic/clinical characteristics

Stratified admission rates for respiratory and GI infections are shown in Appendix Figures 11-22, and stratum-specific IRRs in Appendix Figures 23-26. Although point estimates appeared to vary, there was no evidence of effect modification by any of the characteristics investigated, for either respiratory or GI infection admissions (Appendix Tables 4 and 5).

### Cumulative incidence of first hospitalization

By the age of 1 year, 8-10% of children born in 2017-2019 had been admitted at least once with a primary diagnosis of a respiratory infection depending on birth year (Figure 2A). Inflections in the cumulative incidence curves corresponded to the ages of these children at the onset of the pandemic (e.g. ∼12 months for those born in 2019). For children born in 2020, the cumulative incidence increased more slowly with age, reaching 4% by 12 months. For children born in 2021, cumulative incidence was similar to that for children born pre-pandemic.

For GI infections, cumulative incidence of admission by 12 months was also lowest for those born in 2020 (0.8%, versus 1.0-1.3% for children born pre-pandemic, Figure 2B). For children born in 2021, incidence was similar to those born pre-pandemic.

### Sensitivity analyses

IRRs calculated for overall respiratory and GI infection admissions estimated by 1) including admissions with relevant codes in any position or 2) excluding infants born in London were similar to those obtained in the main analysis (Appendix Table 6). Results did not change in the individual-level analysis of overall respiratory admissions (Appendix Table 7).

## Discussion

In this large, national study, we found substantial reductions in rates of respiratory and GI infection-related hospital admission for children aged <5 years in England, following the onset of the COVID-19 pandemic and implementation of NPIs. These reductions occurred for admissions associated with multiple specific respiratory and GI infections and were particularly marked during the initial lockdown period in spring/summer 2020 but continued into 2021. Rates rebounded to levels higher than usual for the time of year in the spring/summer of 2021. Impacts were greater for respiratory than GI infection admissions and did not vary by sociodemographic or clinical characteristics. Although cumulative incidence of admission with age was low for children born in 2020, children born in 2021 had admission patterns similar to those of children born pre-pandemic.

Our results add to growing evidence, from diverse complementary data sources, showing the impact of COVID-related NPIs on transmission of other respiratory infections [7, 27]. There is also evidence of a decline in childhood vaccination coverage in England during the pandemic [28]. Studies such as ours show the combined effect of the pandemic and response measures on transmission, healthcare-seeking behaviour and, in the longer term, vaccine uptake. Whilst these factors cannot be separated in our analysis, quantifying overall changes in healthcare demand is crucial for planning for the acute and recovery phase of future pandemics, given that NPIs will likely be considered as control measures for respiratory pathogens such as influenza. Ultimately, understanding and planning for these impacts will enhance pandemic preparedness and ensure resilient healthcare systems. Our results are consistent with the large reductions in numbers of admissions for a range of infections recorded in HES for 0-14 year-olds [11], as well as with results from elsewhere [2, 29], and provide additional focus on the youngest age group including vulnerable sub-groups, over a longer period.

There is less published evidence of an effect of the pandemic and NPIs on GI infections in children [9]. Data from England, not stratified by age, show reductions in laboratory isolations of GI pathogens during NPI implementation, which reversed with the lifting of NPIs [27]. In Bavaria, Germany, notification rates for rotavirus, norovirus and salmonellosis were 65%, 85% and 40% lower, respectively during March-December 2020 versus the pre-pandemic yearly average, with similar reductions in 2021 [30].

Acute bronchiolitis is largely caused by respiratory syncytial virus (RSV) [31], circulation of which has been disrupted since the COVID-19 pandemic [2, 32]. Consistent with this, we saw changes to the typical seasonality of respiratory admissions in 2021, particularly for acute bronchiolitis for which admission rates were over 5 times higher in July-September 2021 compared to July-September of pre-pandemic years. Although rates were below pre-pandemic seasonal peaks, this high IRR shows a considerable excess over the rates expected for the time of year, with implications for resource planning; this adds granularity to previous analyses showing increases in annual admission numbers or rates post-pandemic [33]. A national Danish study reported changes in incidence of laboratory-confirmed RSV of similar magnitude amongst 0-17 year-olds, with IRRs of 0.10 (0.09-0.11) and 4.51 (4.41-4.60) comparing the lockdown and post-lockdown periods, respectively, to pre-lockdown [34].

These post-pandemic resurgences may stem from reduced immunity due to lower pathogen exposure (and re-exposure following lifting of NPIs), and interactions between pathogens [9]. RSV is also a cause of croup [35], which may explain the off-season peaks in croup admissions in 2020 and 2021. Admission rates for GI infections were also markedly reduced following introduction of NPIs in our study, broadly comparable to results of a single-centre Danish study (70% decrease in admission rates for GI infections amongst 0-5 year-olds during March-June 2020 compared to the pre-pandemic average) [36].

We did not find statistical evidence that the impact of NPIs (as reflected in calendar period) on respiratory or GI admissions differed according to clinical or sociodemographic characteristics. This is consistent with previous analyses of HES data showing declines in numbers of admissions for children aged <15 years across demographic and clinical groups [11]. Factors not explored here, such as differences between sociodemographic groups in household crowding and access to childcare, may influence the impact of NPIs and should be explored in future studies.

Strengths of the study include the large size and longitudinal nature of the dataset, with almost complete ascertainment of births and hospitalizations in England and good availability of data on sociodemographic and clinical characteristics relevant to healthcare use and infection risk. We compared post-NPI admission rates to those in the corresponding quarters of previous years, accounting for expected seasonal variation which was disrupted by the pandemic.

We could not identify children who emigrated from England, so denominators could be overestimated and rates underestimated, especially for older children; the impact of this is likely small as few young children emigrate (e.g. an estimated ∼6000 0-17 year-olds in 2018 [37]), and travel restrictions limited emigration during the pandemic. Coders entering data into HES follow standardized procedures to code information from patient notes, but recording of this information by clinicians may vary between hospitals and over time, particularly during the pandemic [16]. However, results showed a substantial impact of NPIs for both broad and more specific outcomes. Data were missing for some variables, notably IMD and preterm birth (available for 80% and 78% of children, respectively). Whilst there were no clear differences between those with and without data on preterm birth, children without data on IMD were less likely to be born preterm and to have congenital anomalies (both of which are associated with lower socioeconomic position [38, 39]), and also more likely to be admitted, which might have introduced some selection bias in analyses using this variable. Recording of ethnicity in HES does not always agree with self-reported ethnicity in the UK Census (with lower concordance for non-white ethnicities) [40]. IMD is a measure of area-level, rather than individual-level, deprivation; additionally we did not allow for changes in deprivation since birth. Our main regression analyses treated repeat admissions as independent events, but few children had >1 admission and analysis of respiratory admissions (for which repeat admissions are more common compared to GI infections) allowing for clustering by patient generated almost identical results. Finally, longer-term analyses are needed to fully elucidate the impact of the pandemic on hospital admissions and across the healthcare system (for example, A&E attendances are collected in a separate database [16]).

In conclusion, we demonstrate long-term changes in infection-related hospital admissions for children aged <5 years in England following the onset of the COVID-19 pandemic. These were independent of children’s demographic and clinical characteristics and continued throughout 2021. Our results add further insight into the breadth of impact of the SARS-CoV-2 pandemic, and offer useful evidence on the scale of longer-term consequences which can be used to inform public health planning for future pandemics and their aftermath. Continued monitoring of the current situation is needed to further inform planning for recovery from future pandemics.

## Data Availability

This study uses NHS Hospital Episode Statistics data and was provided within the terms of a data-sharing agreement (DARS-NIC-393510-D6H1D-v9.3) to the researchers by NHS England. The data do not belong to the authors and may not be shared by the authors, except in aggregate form for publication. The data is provided by patients and collected by the NHS as part of their care and support. Data can be obtained by submitting a data request through the NHS England Data Access Request Service.

## Financial support

This work is part of the VERDI project (101045989) which is funded by the European Union. This work is partly supported by the NIHR Great Ormond Street Hospital Biomedical Research Centre. The MRC Clinical Trials Unit at UCL is supported by the Medical Research Council (programme number: MC_UU_00004/03). Views and opinions expressed are however those of the author(s) only and do not necessarily reflect those of the European Union or the European Health and Digital Executive Agency, the NHS, the NIHR or the Department of Health. Neither the European Union nor the granting authority can be held responsible for them. The funders had no role in study design, data collection and analysis, decision to publish, or preparation of the manuscript.

## Declaration of interests

CJ, LW, FA, ML, AJ, CT, IJC and PH had financial support from the European Union (payments to institutions); CT declares a grant from the GOSH Biomedical Research Centre (payments to institution) and board membership of the Penta Fondazione Penta ETS. CJ, AJ and IJC are affiliated to the MRC Clinical Trials Unit at UCL, which receives funding from the Medical Research Council.

## Statement of authors’ contributions

PH conceived the study. FA constructed the birth cohort. CJ analysed the data with substantial input from LW and PH, and drafted the paper. All authors contributed to interpretation of results and critically reviewed and revised the paper.

