## Appendix for "Paediatric infection hospital admissions in England before, during and after the COVID-19 pandemic"

Charlotte Jackson^a^*, Linda Wijlaars^b^, Fariyo Abdullahi^b^, Mengyun Liu^b^, Ali Judd^a,c^, Claire Thorne^b^, Intira Jeannie Collins^a^, Costanza Di Chiara^d^, Pia Hardelid^b,e^

^a^ MRC Clinical Trials Unit at UCL, 90 High Holborn, London WC1V 6LJ, UK

^b^ UCL Great Ormond Street Institute for Child Health, 30 Guilford Street, London WC1N 1EH, UK

^c^ Fondazione Penta ETS, Padua, Italy

^d^ Department for Women's and Children's Health, University of Padova, Via Giustiniani, 3 - 35128 Padua, Italy

^e^ National Institute for Health Research Great Ormond Street Hospital Biomedical Research Centre

*Corresponding author:

Charlotte Jackson, MRC Clinical Trials Unit at UCL, 90 High Holborn, London WC1V 6LJ, UK

+44 (0)20 7670 4806

*List of ICD-10 codes used in the analyses*

We used previously published ICD-10 code lists to identify hospitalizations with respiratory or gastrointestinal infection, and additionally included all codes for seasonal influenza, all sites of tuberculosis disease and viral wheeze in respiratory infections, and nausea and vomiting in GI infections (Appendix Table 1).

*Appendix Table 1: ICD-10 codes considered indicative of respiratory and gastrointestinal infections (adapted from Kuan et al 2019) [1].*

| **Respiratory infections** | A06.5, A15, A16, A17*, A18*, A19*, A20.2, A21.2, A22.1, A31.0, A36.0, A36.1, A36.2, A37, A42.0, A43.0, A54.5, A56.4  B01.2, B05.2, B05.3, B25.0, B27, B37.1, B38.0, B38.1, B38.2, B39.0, B39.1, B39.2, B40.0, B40.1, B40.2, B41.0, B42.0, B44.0, B44.1, B44.2, B45.0, B46.0, B58.3, B59, B67.1, B87.3, B87.4  H60, H62.0, H62.1, H62.2, H62.3, H62.4, H65, H66, H67, H70, H73.0, H73.1, H75.0  J00, J01, J02, J03, J04, J05, J06, J10*, J11*, J12, J13, J14, J15, J16, J17, J18, J20, J21, J22, J34.0, J36, J37, J39.0, J39.1, J44.0, J44.1, J65, J85.0, J85.1, J85.2, J86  N740*, N741*  P23  B34.9 (primary) + R06.2 (secondary)* |
| --- | --- |
| **Gastrointestinal infections** | A00, A01, A02.0, A03, A04, A05, A06.0, A06.1, A06.2, A06.3, A07, A08, A09, A18.3, A21.3, A22.2  B05.4, B46.2, B78.0, B81, B82, B98.0  K23.0, K23.1, K63.0, K93.0  R11* |

*Code not included in original list

*Appendix Table 2: Characteristics of children with and without data on IMD.*

|  | No IMD | Has IMD |
| --- | --- | --- |
|  | 1,204,809 | 4,779,452 |
| Sex (n = 5,982,720) |  |  |
| Male | 600,467 (49.9%) | 2,468,910 (51.7%) |
| Female | 603,653 (50.1%) | 2,309,690 (48.3%) |
| Congenital anomalies (n = 5,984,261) |  |  |
| No | 1,187,939 (98.6%) | 4,601,977 (96.3%) |
| Yes | 16,870 (1.4%) | 177,475 (3.7%) |
| Ethnicity (n = 5,544,132) |  |  |
| White | 800,582 (72.8%) | 3,321,514 (74.7%) |
| Asian or Asian British | 142,059 (12.9%) | 533,461 (12.0%) |
| Black, Black British, Caribbean or African | 60,298 (5.5%) | 214,579 (4.8%) |
| Mixed or multiple ethnic groups | 68,324 (6.2%) | 258,964 (5.8%) |
| Any other ethnic group | 29,147 (2.6%) | 115,204 (2.6%) |
| Preterm (n = 4,696,490) |  |  |
| Term | 829,164 (95.6%) | 3,562,280 (93.0%) |
| Preterm | 37,826 (4.4%) | 267,220 (7.0%) |
| Maternal age (years) (n = 4,203,342) | 30 [25 - 34] | 30 [26 - 34] |
| Mother born outside UK (n = 3,495,873) |  |  |
| UK | 285,786 (66.7%) | 2,190,550 (71.4%) |
| Non UK | 142,473 (33.3%) | 877,064 (28.6%) |
| Total number of respiratory admissions (n = 5,984,261) (median [IQR]) | 0 [0 - 0] | 0 [0 - 0] |
| Total number of respiratory admissions (n = 5,984,261) |  |  |
| 0 | 1,160,182 (96.3%) | 4,272,574 (89.4%) |
| 1 | 36,339 (3.0%) | 375,114 (7.8%) |
| >1 | 8,288 (0.7%) | 131,764 (2.8%) |
| Total number of GI admissions (n = 5,984,261) (median [IQR]) | 0 [0 - 0] | 0 [0 - 0] |
| Total number of GI admissions (n = 5,984,261) |  |  |
| 0 | 1,197,097 (99.4%) | 4,685,322 (98.0%) |
| 1 | 7,411 (0.6%) | 86,414 (1.8%) |
| >1 | 301 (0.0%) | 7,716 (0.2%) |

*Appendix Table 3: Characteristics of children with and without data on gestational age (GA) at birth.*

|  | No GA | Has GA |
| --- | --- | --- |
|  | 1,287,771 | 4,696,490 |
| Sex (n = 5,982,720) |  |  |
| Male | 659,950 (51.3%) | 2,409,427 (51.3%) |
| Female | 627,377 (48.7%) | 2,285,966 (48.7%) |
| Congenital anomalies (n = 5,984,261) |  |  |
| No | 1,241,617 (96.4%) | 4,548,299 (96.8%) |
| Yes | 46,154 (3.6%) | 148,191 (3.2%) |
| Ethnicity (n = 5,544,132) |  |  |
| White | 837,474 (73.9%) | 3,284,622 (74.5%) |
| Asian or Asian British | 128,920 (11.4%) | 546,600 (12.4%) |
| Black, Black British, Caribbean or African | 56,587 (5.0%) | 218,290 (4.9%) |
| Mixed or multiple ethnic groups | 74,660 (6.6%) | 252,628 (5.7%) |
| Any other ethnic group | 35,555 (3.1%) | 108,796 (2.5%) |
| Maternal age (years) (n = 4,203,342) | 30 [26 - 34] | 30 [26 - 34] |
| Mother born outside UK (n = 3,495,873) |  |  |
| UK | 508,843 (69.3%) | 1,967,493 (71.2%) |
| Non UK | 225,114 (30.7%) | 794,423 (28.8%) |
| IMD quintile (n = 4,779,452) |  |  |
| Quintile 1 (most deprived) | 239,195 (25.2%) | 1,077,311 (28.1%) |
| Quintile 2 | 225,749 (23.8%) | 843,677 (22.0%) |
| Quintile 3 | 187,869 (19.8%) | 710,184 (18.5%) |
| Quintile 4 | 160,022 (16.8%) | 614,976 (16.1%) |
| Quintile 5 (least deprived) | 137,117 (14.4%) | 583,352 (15.2%) |
| Total number of respiratory admissions (n = 5,984,261) (median [IQR]) | 0 [0 - 0] | 0 [0 - 0] |
| Total number of respiratory admissions (n = 5,984,261) |  |  |
| 0 | 1,172,071 (91.0%) | 4,260,685 (90.7%) |
| 1 | 86,588 (6.7%) | 324,865 (6.9%) |
| >1 | 29,112 (2.3%) | 110,940 (2.4%) |
| Total number of GI admissions (n = 5,984,261) (median [IQR]) | 0 [0 - 0] | 0 [0 - 0] |
| Total number of GI admissions (n = 5,984,261) |  |  |
| 0 | 1,266,471 (98.3%) | 4,615,948 (98.3%) |
| 1 | 19,660 (1.5%) | 74,165 (1.6%) |
| >1 | 1,640 (0.1%) | 6,377 (0.1%) |

*Appendix Table 4: Difference in AIC obtained from models for respiratory admissions including and not including an interaction between calendar period and demographic / clinical characteristics.*

| Quarter | AIC (with interaction) | | | | | |
| --- | --- | --- | --- | --- | --- | --- |
|  | Sex | Age group | Congenital anomalies | IMD quintile | Preterm | Ethnicity |
| Apr-June 2020 | 61.69 | 74.48 | 72.76 | 188.85 | 67.8 | 179.86 |
| Jul-Sept 2020 | 57.46 | 141.62 | 59.15 | 179.2 | 51.03 | 193.79 |
| Oct-Dec 2020 | 71.79 | 42.85 | 54.13 | 221.53 | 80.49 | 211.06 |
| Jan-Mar 2021 | 76.1 | 275.65 | 53.43 | 196.92 | 60.49 | 183.32 |
| Apr-Jun 2021 | 63.57 | 80.19 | 74.57 | 196.42 | 69.59 | 188.1 |
| Jul-Sept 2021 | 58.71 | 145.19 | 60.25 | 183.9 | 52.16 | 198 |
| Oct-Dec 2021 | 72.81 | 45.86 | 55.11 | 225.18 | 81.52 | 214.23 |

*Appendix Table 5: Difference in AIC obtained from models for gastrointestinal admissions including and not including an interaction between calendar period and demographic / clinical characteristics.*

| Quarter | AIC (with interaction) | | | | | |
| --- | --- | --- | --- | --- | --- | --- |
|  | Sex | Age group | Congenital anomalies | IMD quintile | Preterm | Ethnicity |
| Apr-June 2020 | 40.51 | 214.23 | 32 | 165.09 | 37.73 | 142.26 |
| Jul-Sept 2020 | 40.47 | 130.61 | 37.36 | 157.36 | 35.31 | 146.19 |
| Oct-Dec 2020 | 45.51 | 124.89 | 36.44 | 157.39 | 40.59 | 135.77 |
| Jan-Mar 2021 | 48.43 | 628.7 | 41.68 | 152.73 | 45.64 | 153.52 |
| Apr-Jun 2021 | 41.15 | 216.48 | 32.4 | 167.43 | 38.1 | 145.54 |
| Jul-Sept 2021 | 41.43 | 133.96 | 37.99 | 161.06 | 36.05 | 149.44 |
| Oct-Dec 2021 | 46.56 | 128.6 | 37.17 | 161.24 | 41.45 | 139.42 |

*Appendix Table 6: Incidence rate ratios comparing quarterly admission rates indicating respiratory or gastrointestinal infections with corresponding pre-pandemic quarters obtained from sensitivity analysis using admissions with relevant diagnostic codes in any position, or excluding children born in London*

|  | **Respiratory infections** | | | | **Gastrointestinal infections** | | | |
| --- | --- | --- | --- | --- | --- | --- | --- | --- |
|  | Code in any position | | Excluding children born in London | | Code in any position | | Excluding children born in London | |
|  | IRR | 99% CI | IRR | 99% CI | IRR | 99% CI | IRR | 99% CI |
| Apr-Jun 2020 | 0.18 | (0.18-0.19) | 0.18 | (0.17-0.19) | 0.37 | (0.35-0.38) | 0.30 | (0.28-0.32) |
| Jul-Sep 2020 | 0.53 | (0.51-0.54) | 0.53 | (0.52-0.54) | 0.50 | (0.48-0.53) | 0.41 | (0.38-0.43) |
| Oct-Dec 2020 | 0.34 | (0.33-0.34) | 0.33 | (0.33-0.34) | 0.41 | (0.39-0.43) | 0.35 | (0.33-0.37) |
| Jan-Mar 2021 | 0.30 | (0.29-0.30) | 0.30 | (0.29-0.31) | 0.44 | (0.43-0.46) | 0.36 | (0.34-0.39) |
| Apr-Jun 2021 | 1.16 | (1.15-1.18) | 1.21 | (1.19-1.23) | 0.68 | (0.66-0.70) | 0.57 | (0.54-0.60) |
| Jul-Sep 2021 | 1.77 | (1.74-1.79) | 1.81 | (1.78-1.83) | 1.13 | (1.09-1.16) | 1.14 | (1.10-1.19) |
| Oct-Dec 2021 | 0.91 | (0.90-0.92) | 0.92 | (0.91-0.93) | 0.98 | (0.95-1.01) | 1.04 | (1.00-1.08) |

*Appendix Table 7:* *Incidence rate ratios comparing quarterly admission rates with primary diagnostic codes indicating respiratory infections with corresponding pre-pandemic quarters, based on individual-level data and accounting for clustering by patient.*

|  | IRR | (99% CI) |
| --- | --- | --- |
| Quarter |  |  |
| April-June 2020 | 0.17 | (0.16-0.18) |
| July-September 2020 | 0.52 | (0.51-0.53) |
| October-December 2020 | 0.34 | (0.33-0.34) |
| January-March 2021 | 0.29 | (0.28-0.29) |
| April-June 2021 | 1.18 | (1.16-1.20) |
| July-September 2021 | 1.79 | (1.76-1.81) |
| October-December 2021 | 0.90 | (0.89-0.91) |

*Appendix Figure 1: Rates of hospital admission in children aged <5 years with a primary diagnostic code indicating acute bronchiolitis (ICD-10 code J21), England, January 2017 to January 2022.*

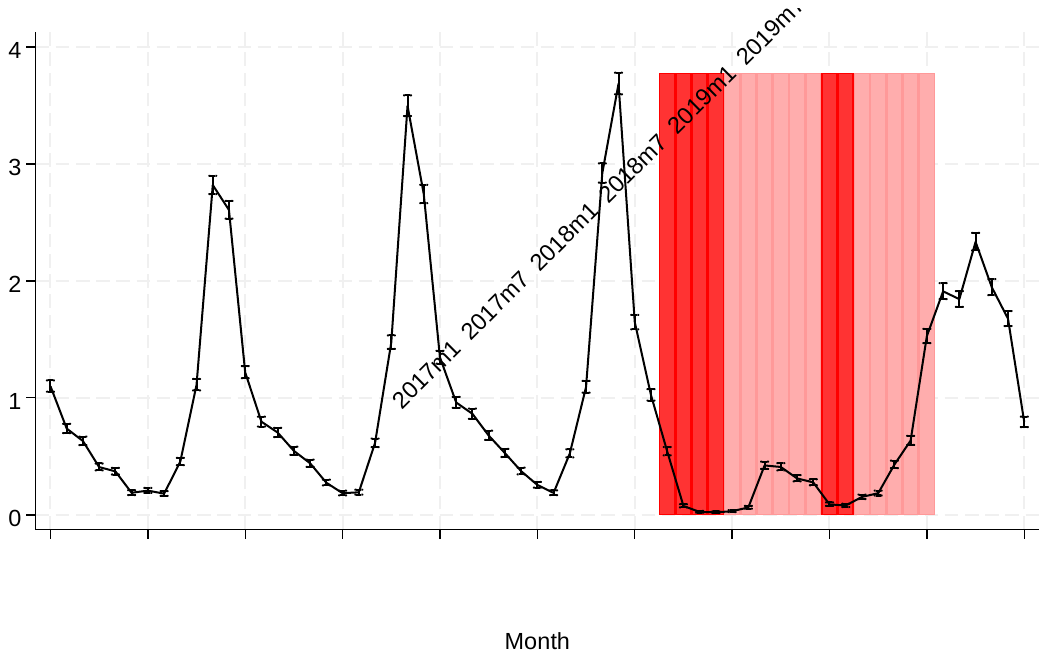

*Appendix Figure 2: Rates of hospital admission in children aged <5 years with a primary diagnostic code indicating acute upper respiratory infections of multiple and unspecified sites (ICD-10 code J06), England, January 2017 to January 2022.*

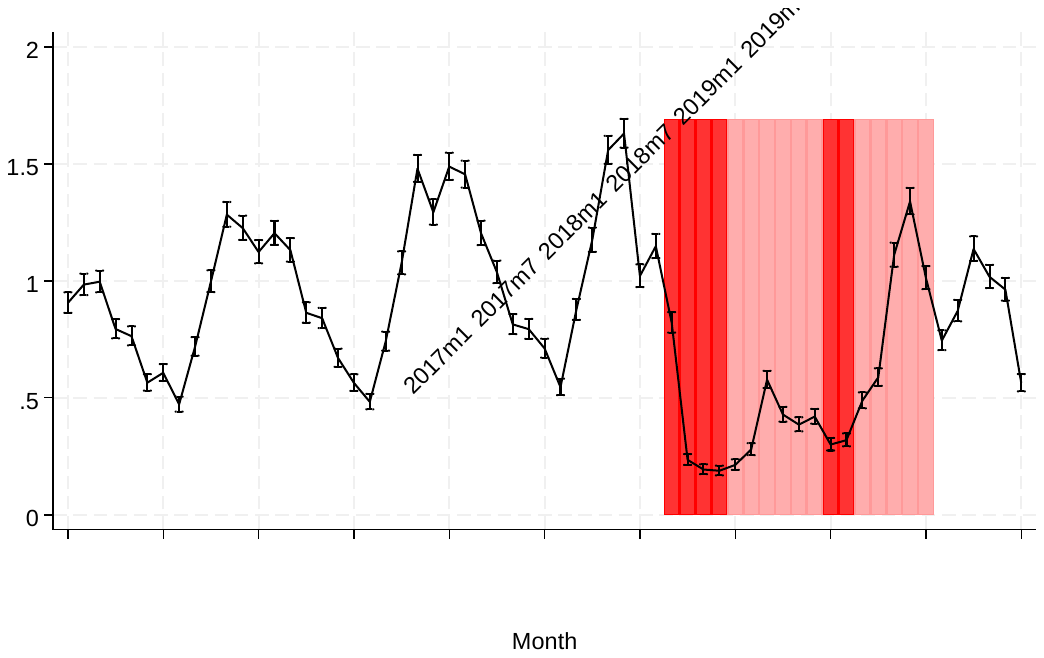

*Appendix Figure 3: Rates of hospital admission in children aged <5 years with a primary diagnostic code indicating unspecified acute lower respiratory infection (ICD-10 code J22), England, January 2017 to January 2022.*

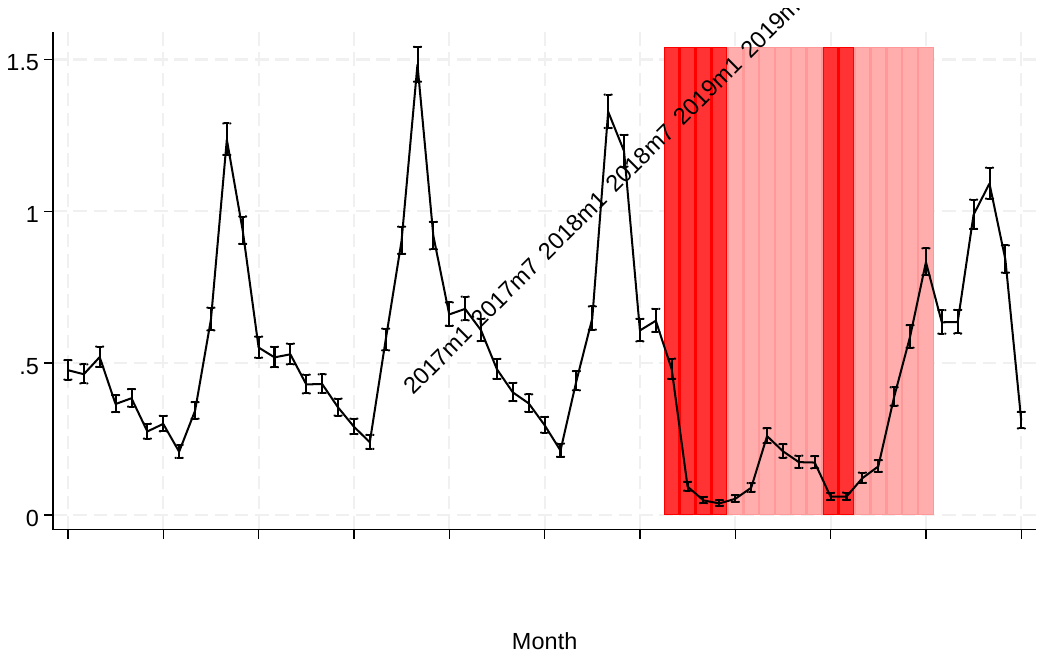

*Appendix Figure 4: Rates of hospital admission in children aged <5 years with a primary diagnostic code indicating acute tonsillitis (ICD-10 code J03), England, January 2017 to January 2022.*

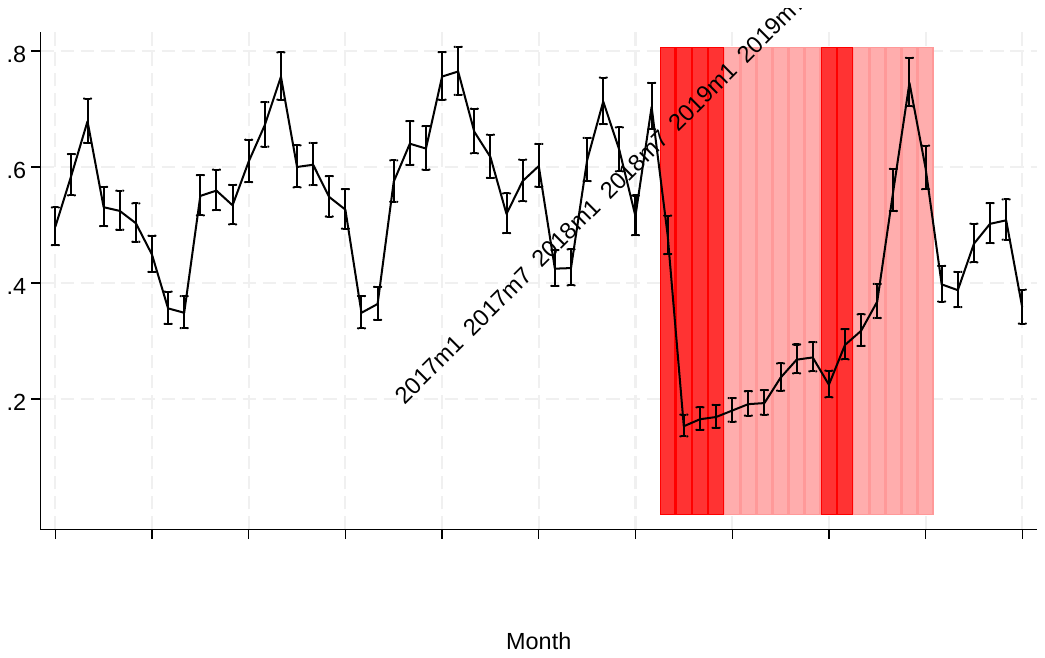

*Appendix Figure 5: Rates of hospital admission in children aged <5 years with a primary diagnostic code indicating acute obstructive laryngitis [croup] and epiglottitis (ICD-10 code J05), England, January 2017 to January 2022.*

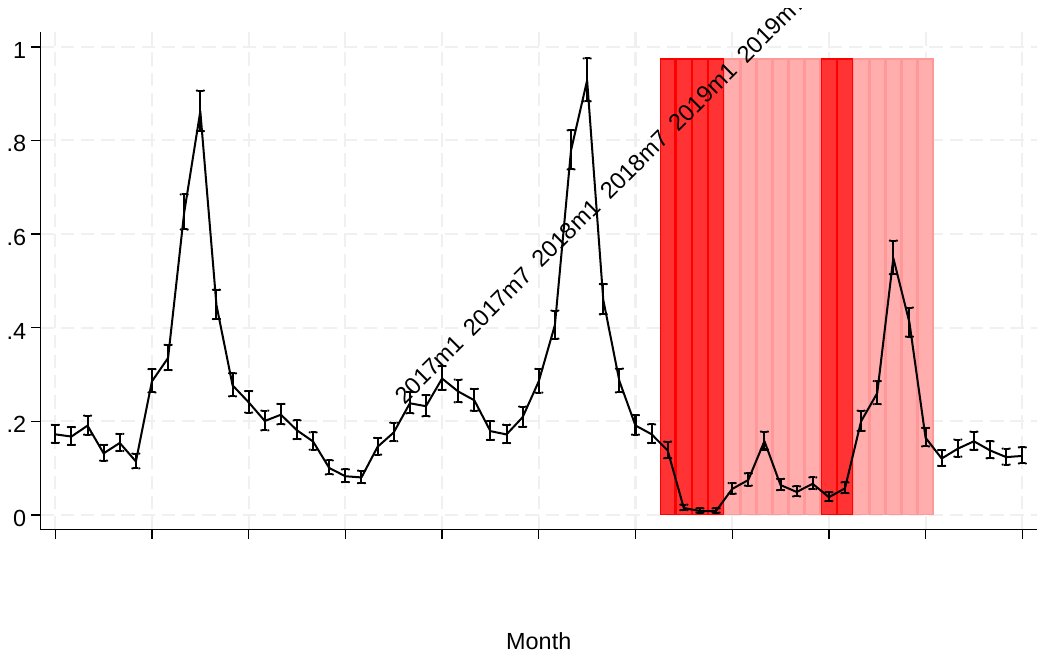

*Appendix Figure 6: Rates of hospital admission in children aged <5 years with diagnostic codes indicating viral wheeze (ICD-10 code B34.9 and R06.2 as primary and secondary diagnoses, respectively), England, January 2017 to January 2022.*

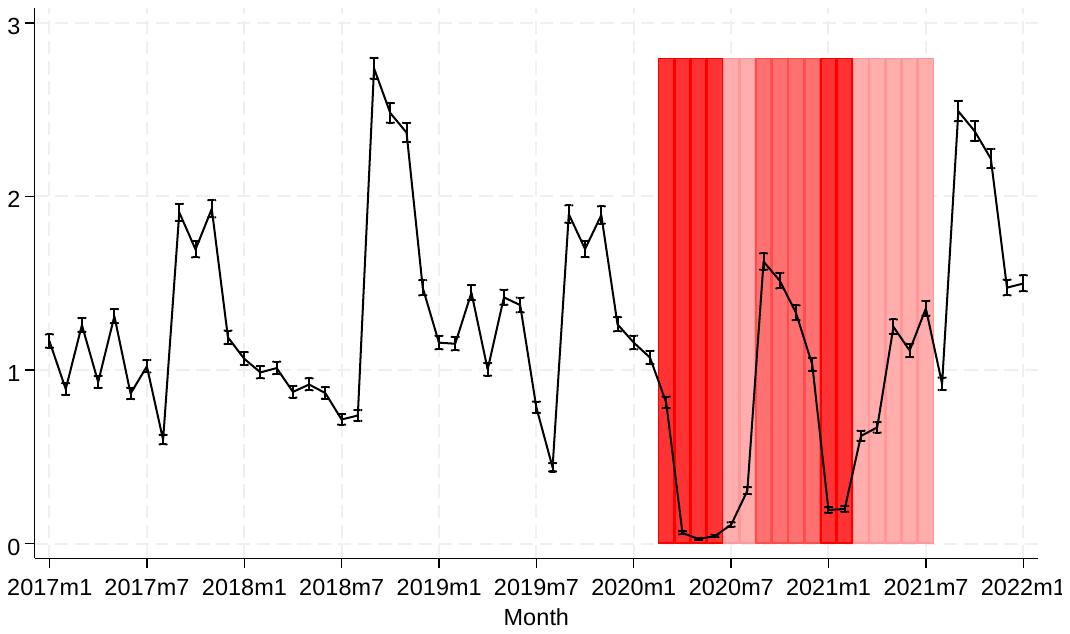

*Appendix Figure 7: Rates of hospital admission in children aged <5 years with a primary diagnostic code indicating viral and other specified intestinal infections (ICD-10 code A08), England, January 2017 to January 2022.*

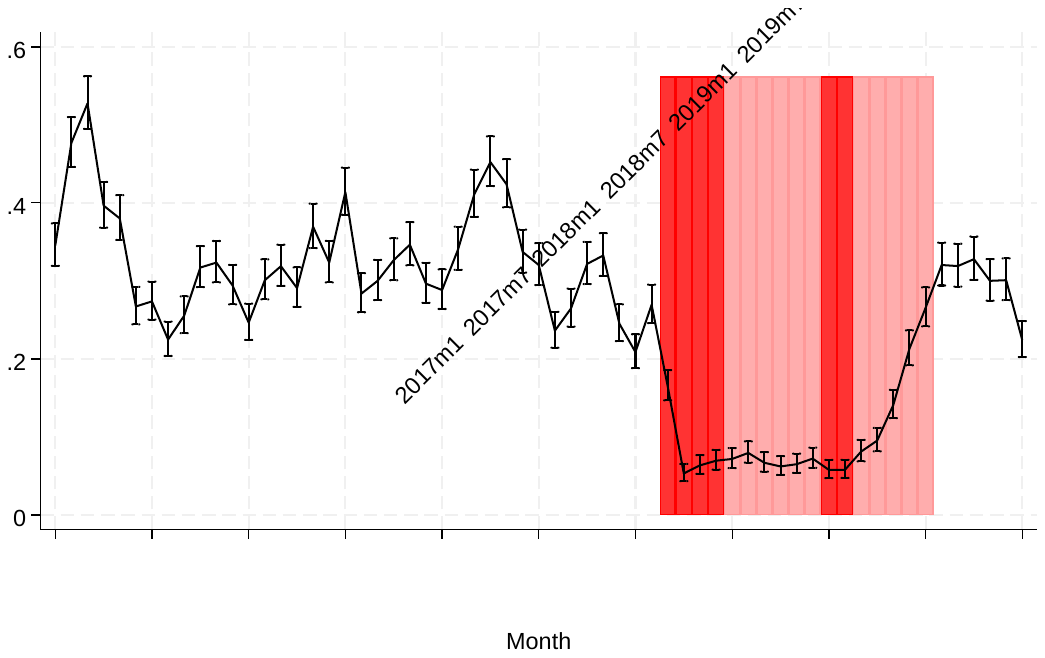

*Appendix Figure 8: Rates of hospital admission in children aged <5 years with a primary diagnostic code indicating other gastroenteritis and colitis of infectious and unspecified origin* *(ICD-10 code A09), England, January 2017 to January 2022.*

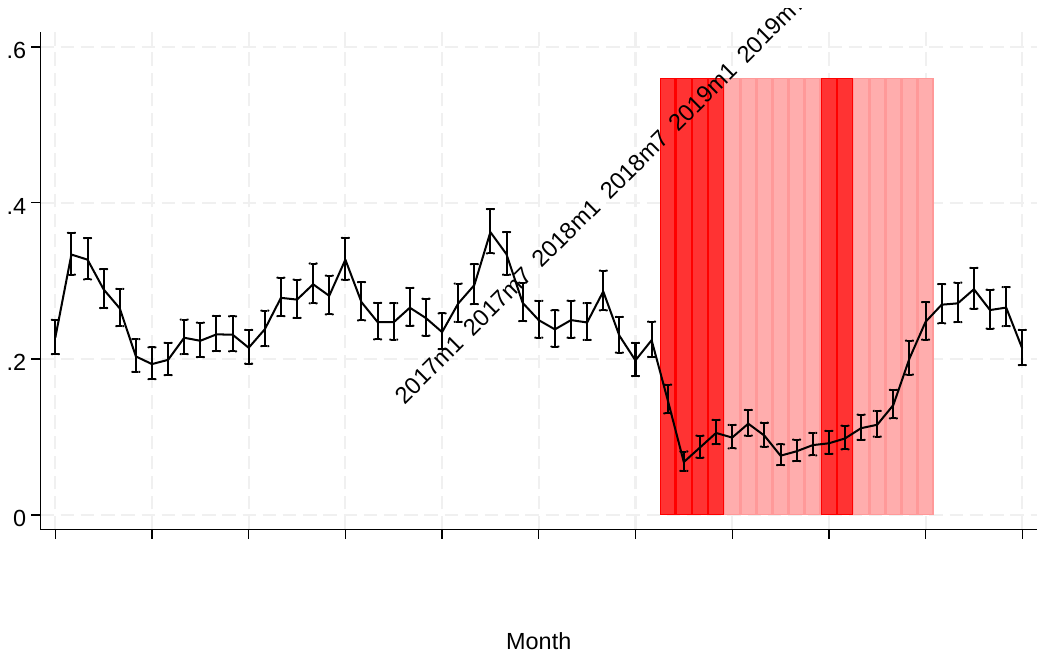

*Appendix Figure 9: Rates of hospital admission in children aged <5 years with a primary diagnostic code indicating other salmonella infections* *(ICD-10 code A02), England, January 2017 to January 2022.*

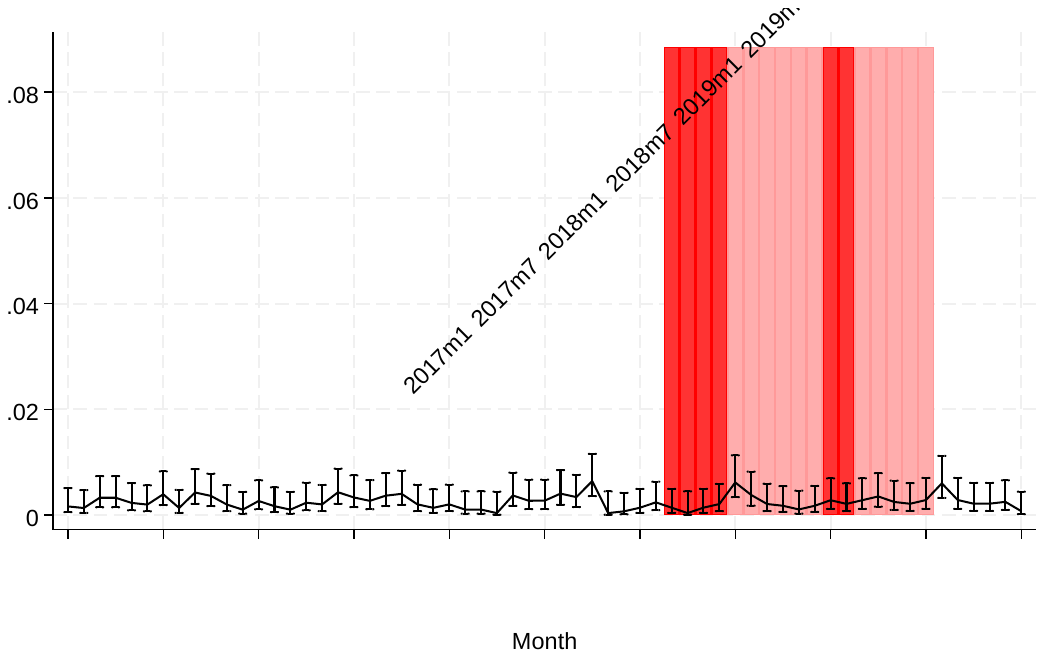

*Appendix Figure 10: Rates of hospital admission in children aged <5 years with a primary diagnostic code indicating nausea and vomiting* *(ICD-10 code R11), England, January 2017 to January 2022.*

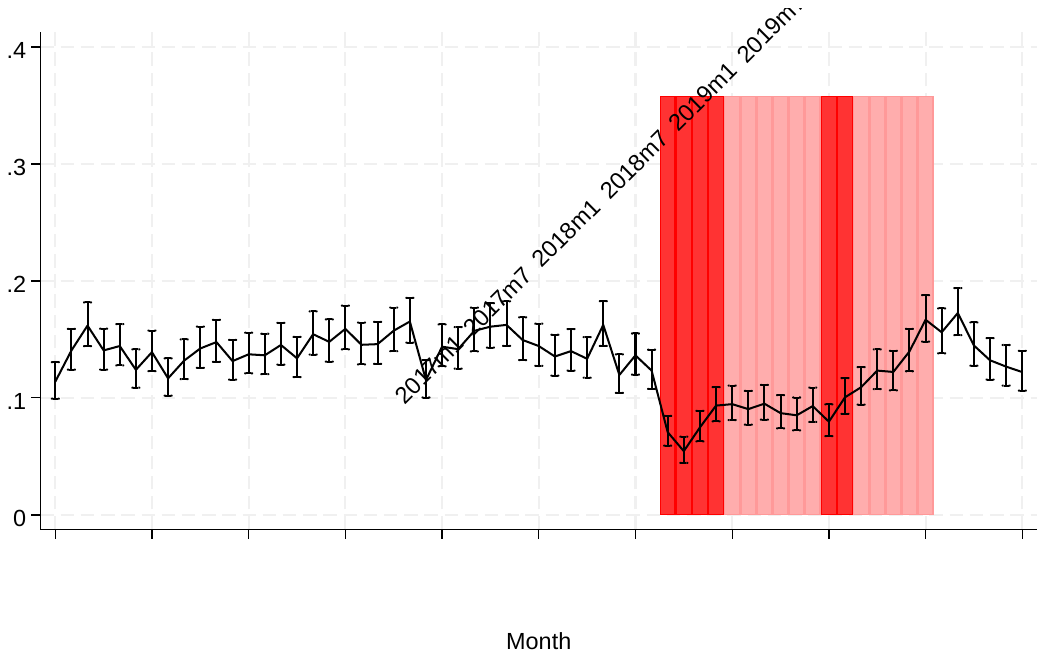

*Appendix Figure 11: Rates of hospital admission in children aged <5 years with a primary diagnostic code indicating a respiratory infection, by sex, England, January 2017 to January 2022.*

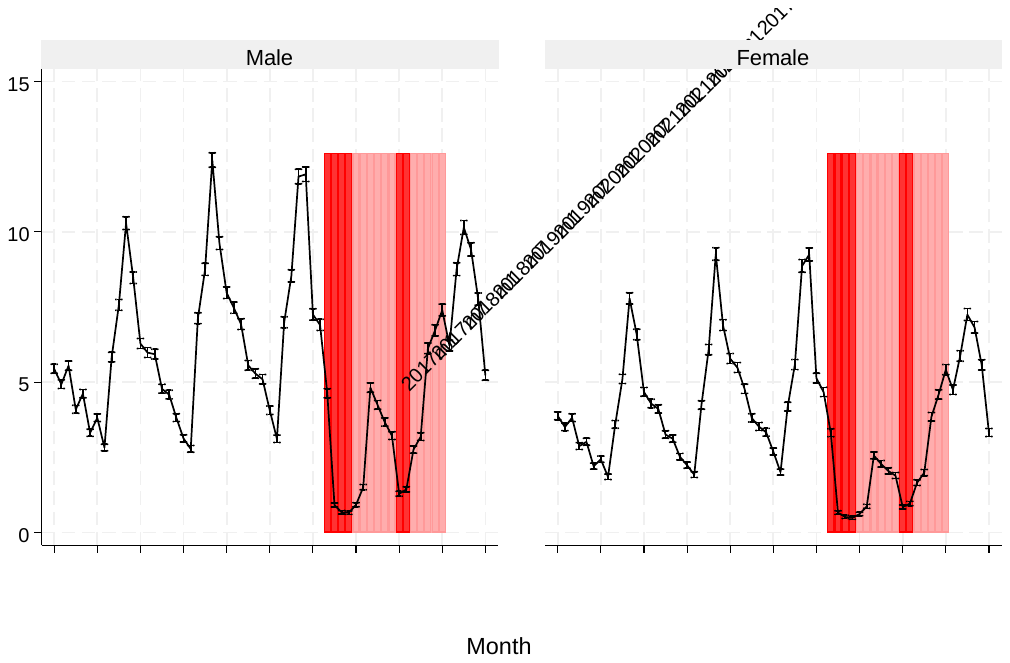

*Appendix Figure 12: Rates of hospital admission in children aged <5 years with a primary diagnostic code indicating a respiratory infection, by age group, England, January 2017 to January 2022.*

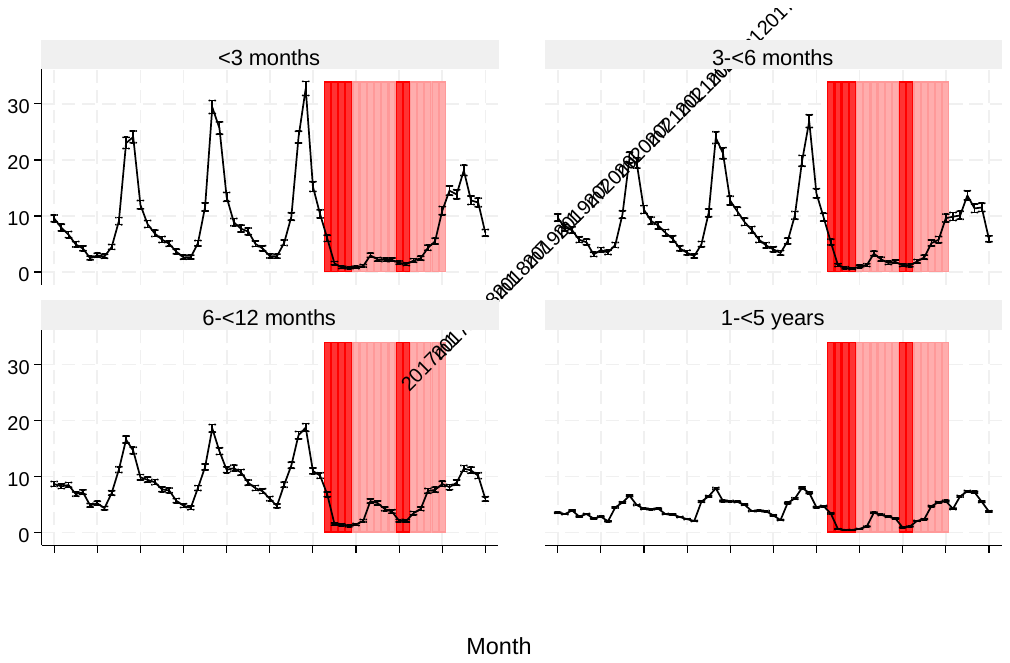

*Appendix Figure 13: Rates of hospital admission in children aged <5 years with a primary diagnostic code indicating a respiratory infection, by presence of congenital anomalies (CA), England, January 2017 to January 2022.*

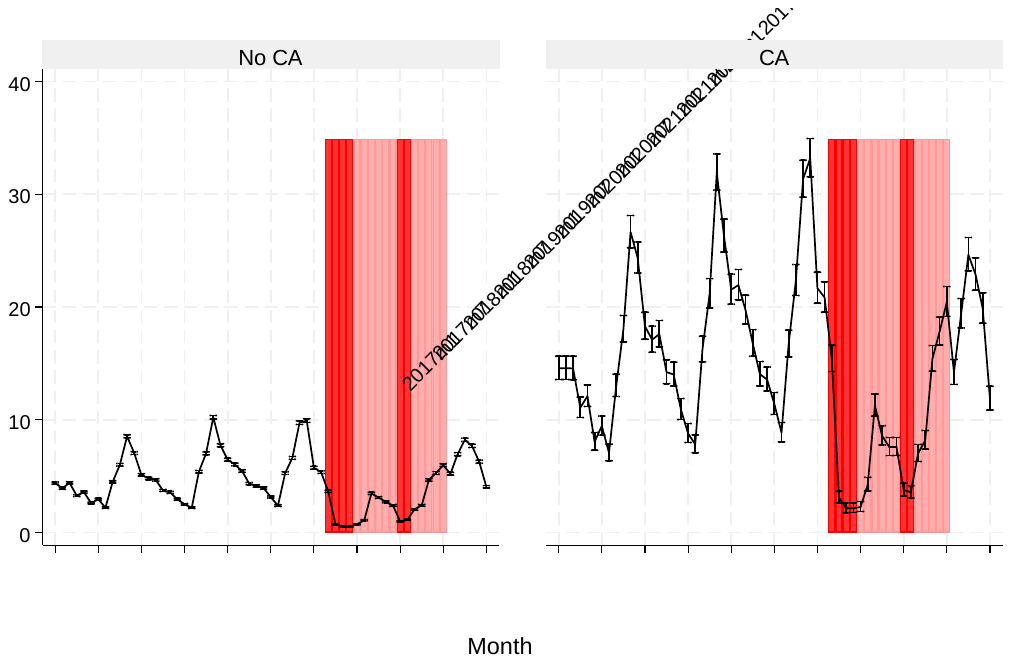

*Appendix Figure 14: Rates of hospital admission in children aged <5 years with a primary diagnostic code indicating a respiratory infection, by preterm birth, England, January 2017 to January 2022.*

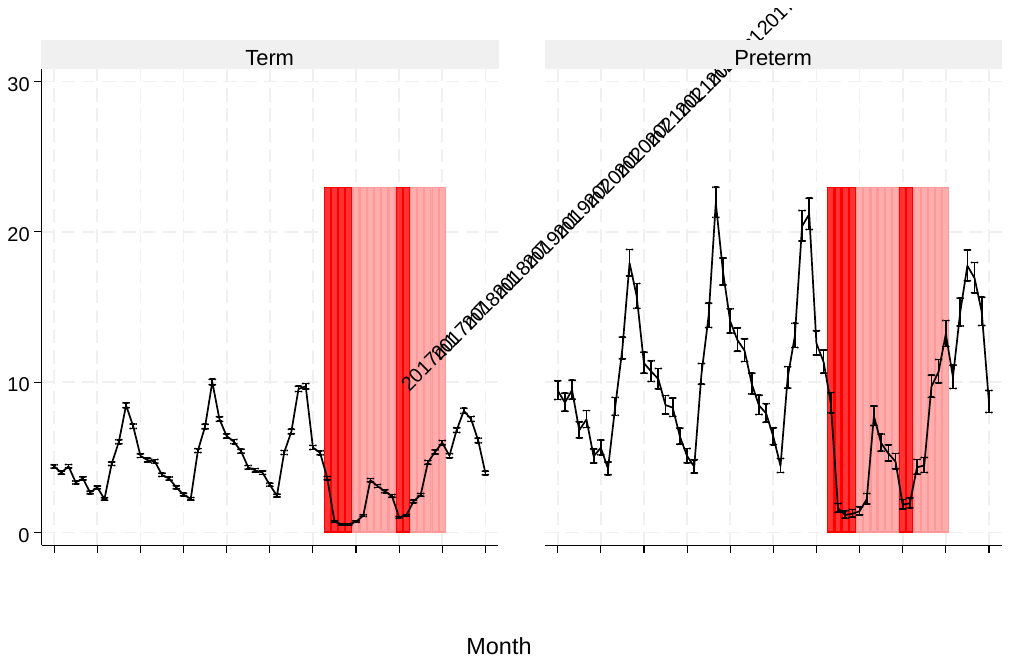

*Appendix Figure 15: Rates of hospital admission in children aged <5 years with a primary diagnostic code indicating a respiratory infection, by IMD quintile, England, January 2017 to January 2022.*

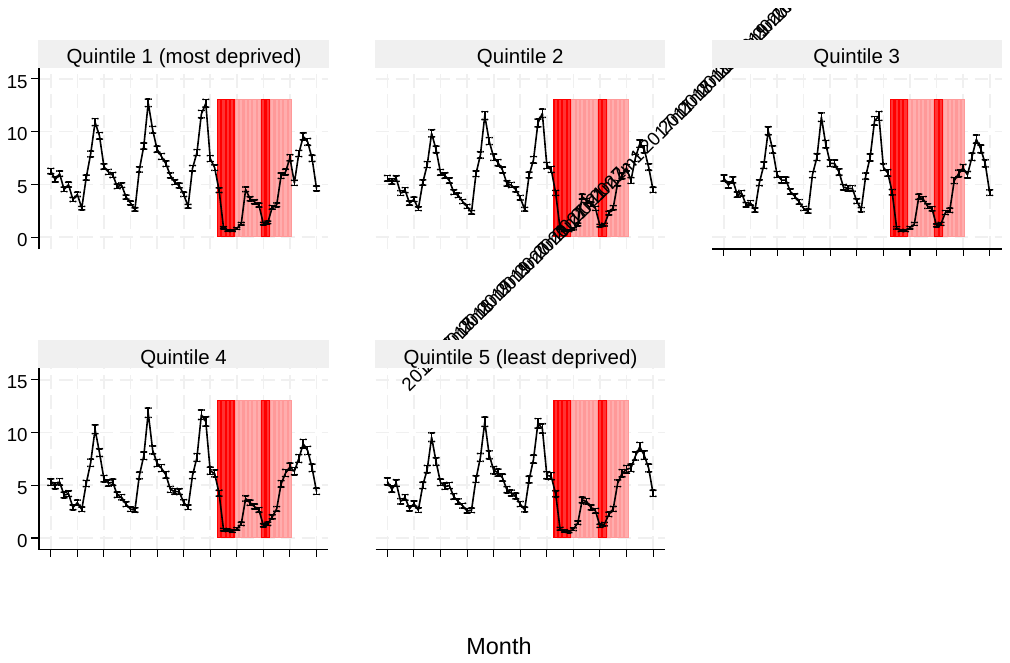

*Appendix Figure 16: Rates of hospital admission in children aged <5 years with a primary diagnostic code indicating a respiratory infection, by ethnicity, England, January 2017 to January 2022.*

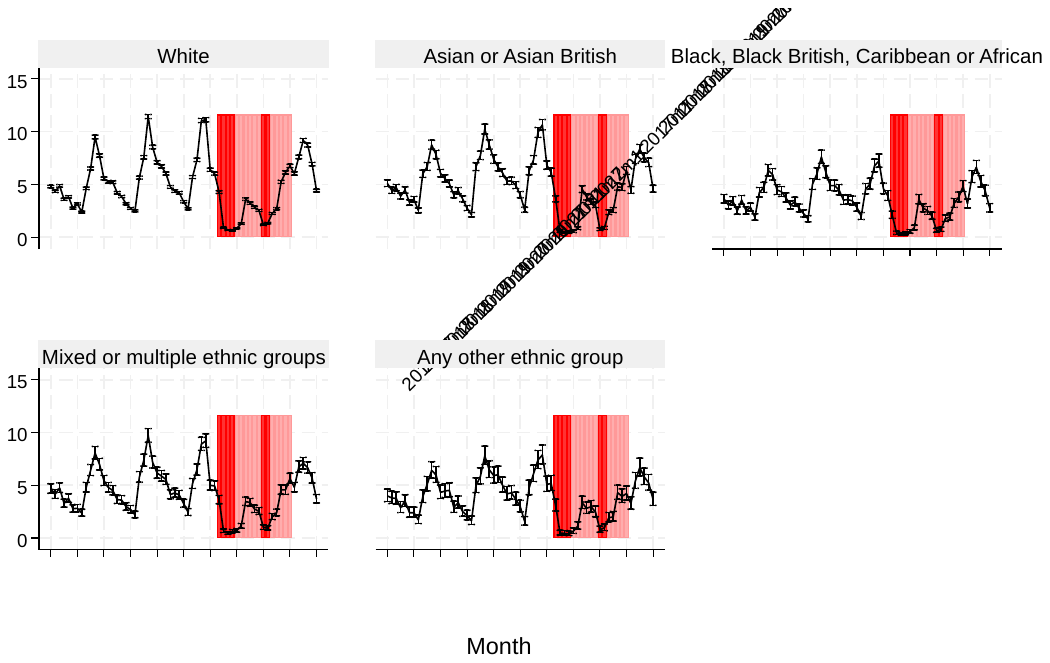

*Appendix Figure 17: Rates of hospital admission in children aged <5 years with a primary diagnostic code indicating a gastrointestinal infection, by sex, England, January 2017 to January 2022.*

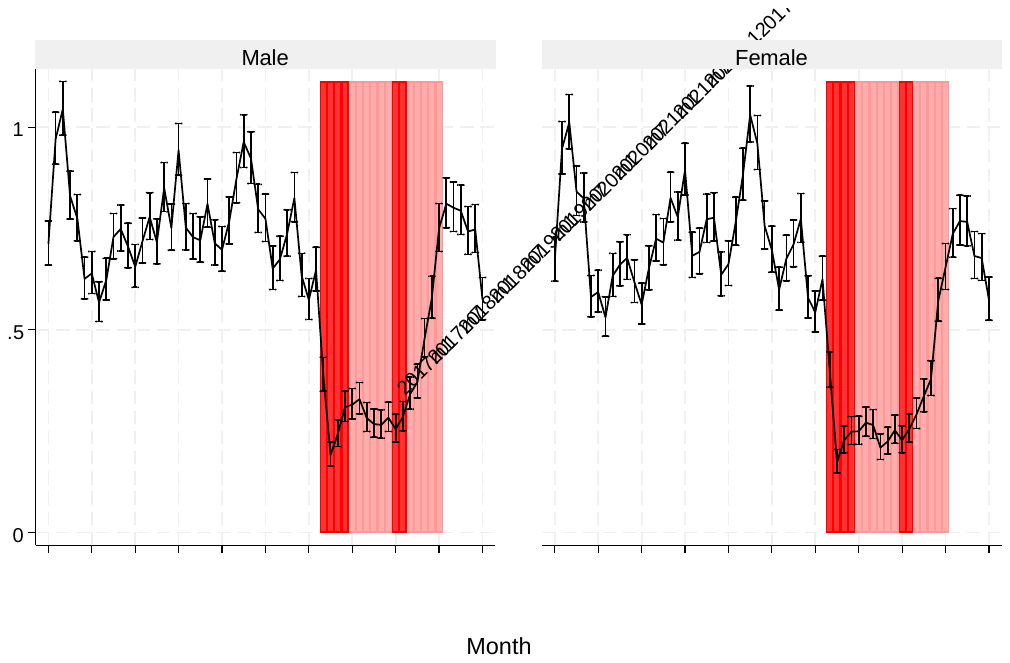

*Appendix Figure 18: Rates of hospital admission in children aged <5 years with a primary diagnostic code indicating a gastrointestinal infection, by age group, England, January 2017 to January 2022.*

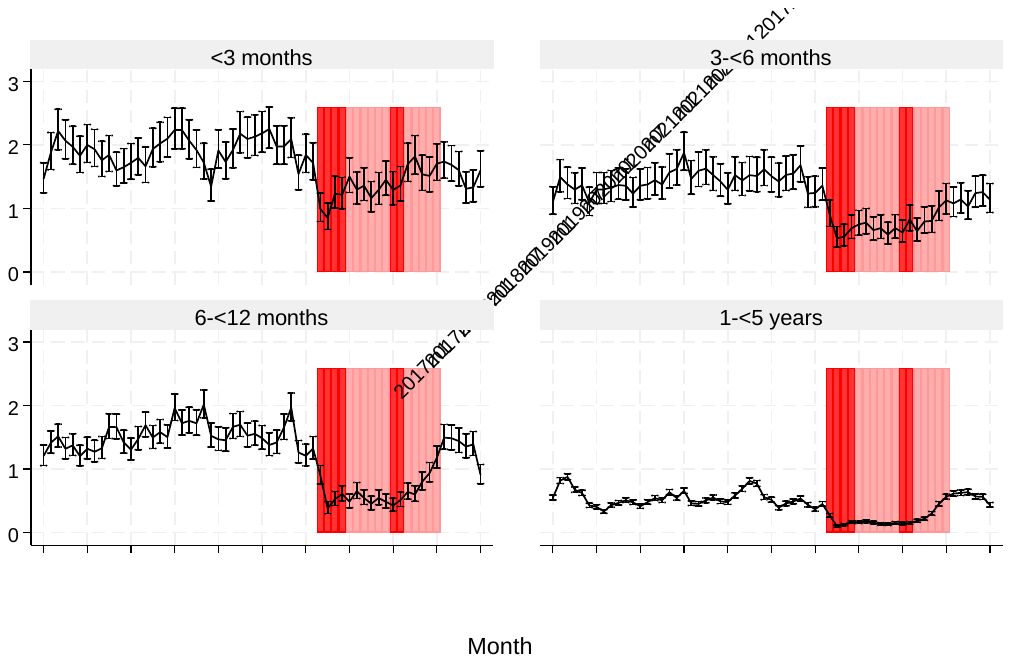

*Appendix Figure 19: Rates of hospital admission in children aged <5 years with a primary diagnostic code indicating a gastrointestinal infection, by presence of congenital anomalies (CA), England, January 2017 to January 2022.*

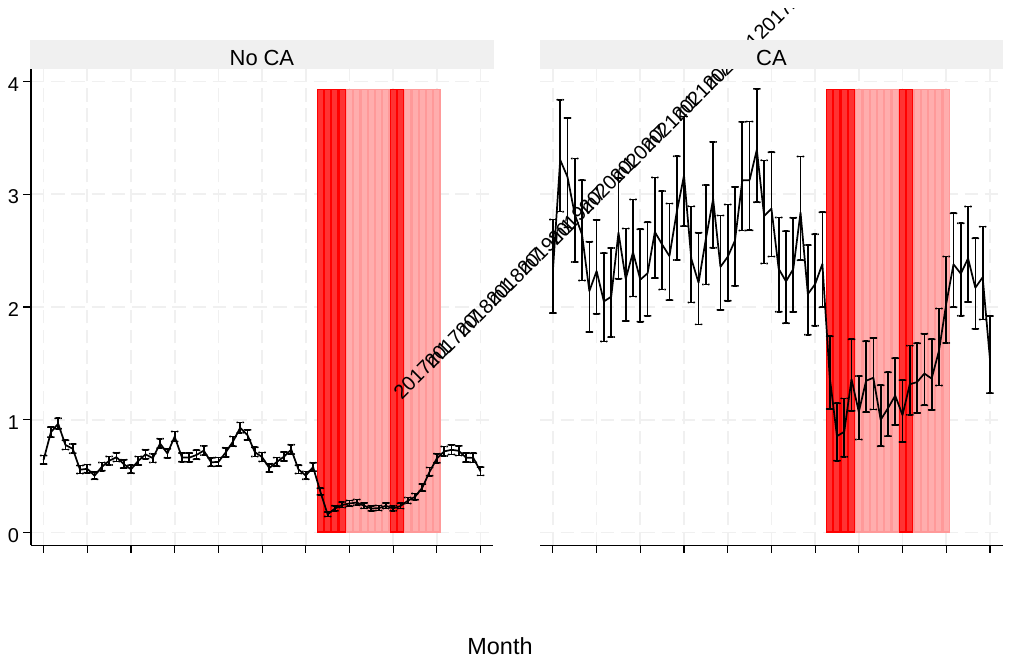

*Appendix Figure 20: Rates of hospital admission in children aged <5 years with a primary diagnostic code indicating a gastrointestinal infection, by preterm birth, England, January 2017 to January 2022.*

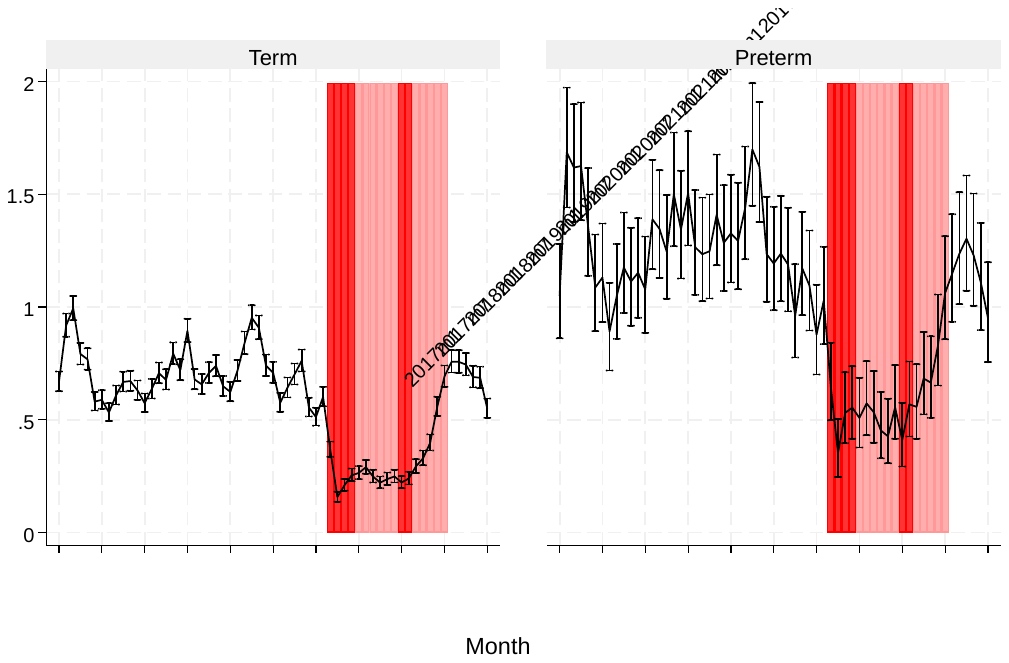

*Appendix Figure 21: Rates of hospital admission in children aged <5 years with a primary diagnostic code indicating a gastrointestinal infection, by IMD quintile, England, January 2017 to January 2022.*

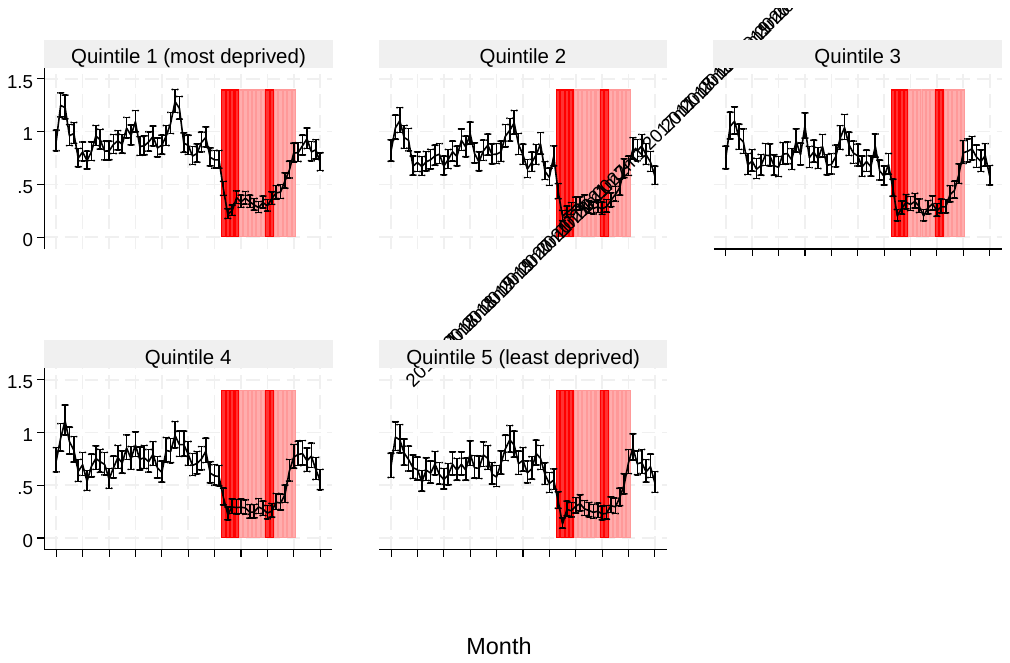

*Appendix Figure 22: Rates of hospital admission in children aged <5 years with a primary diagnostic code indicating a gastrointestinal infection, by ethnicity, England, January 2017 to January 2022.*

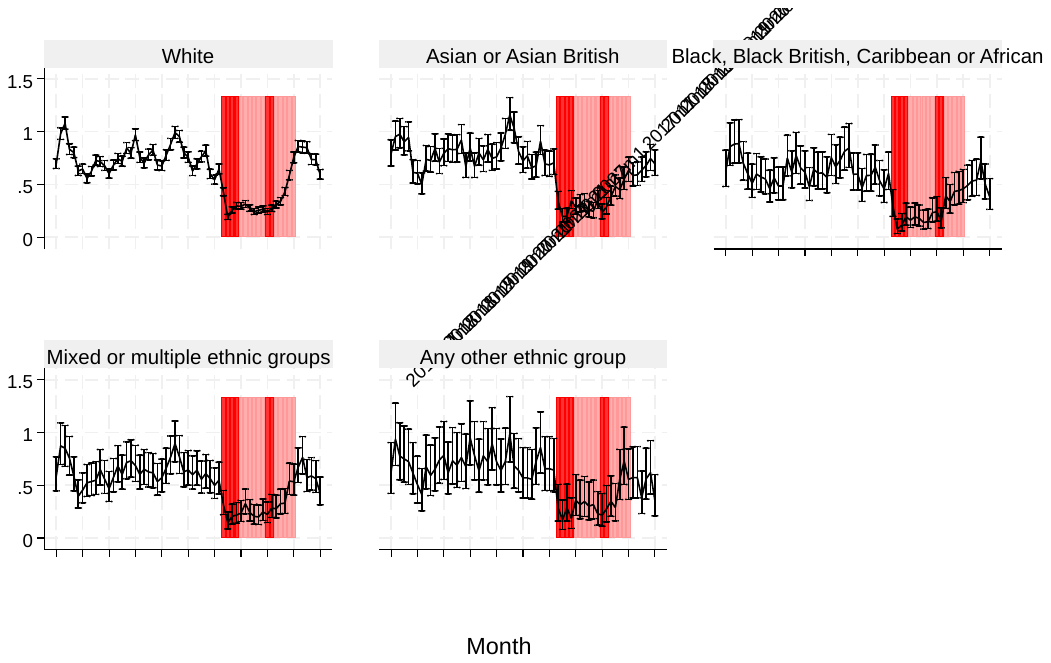

*Appendix Figure 23: Incidence rate ratios comparing quarterly respiratory infection admission rates to corresponding pre-pandemic quarters, by sex, age, area-level deprivation quintile and ethnicity.*

| 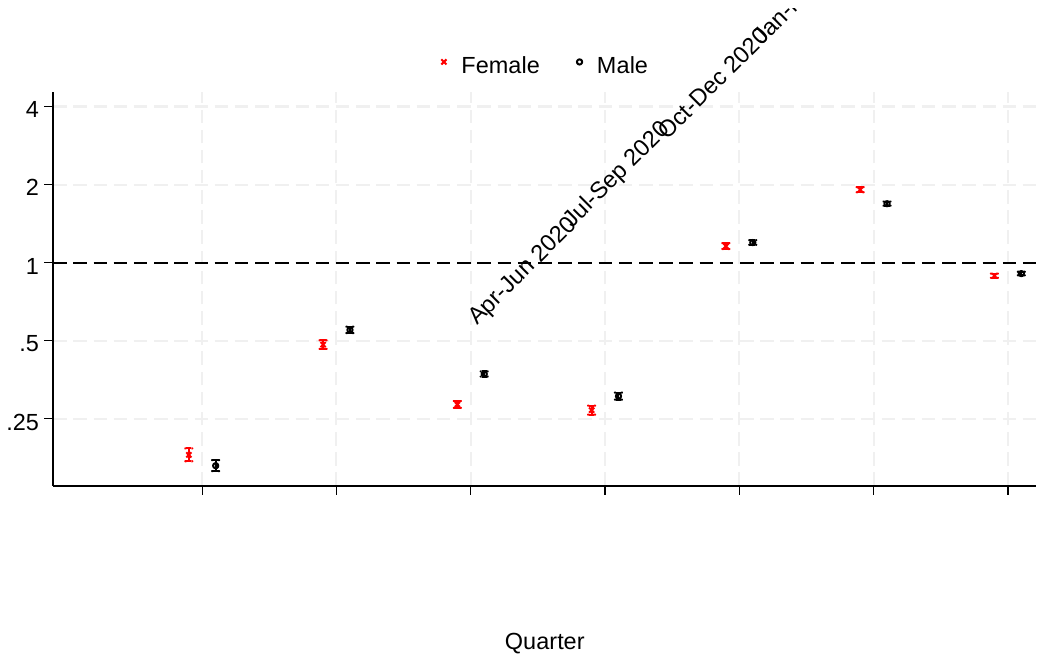 | 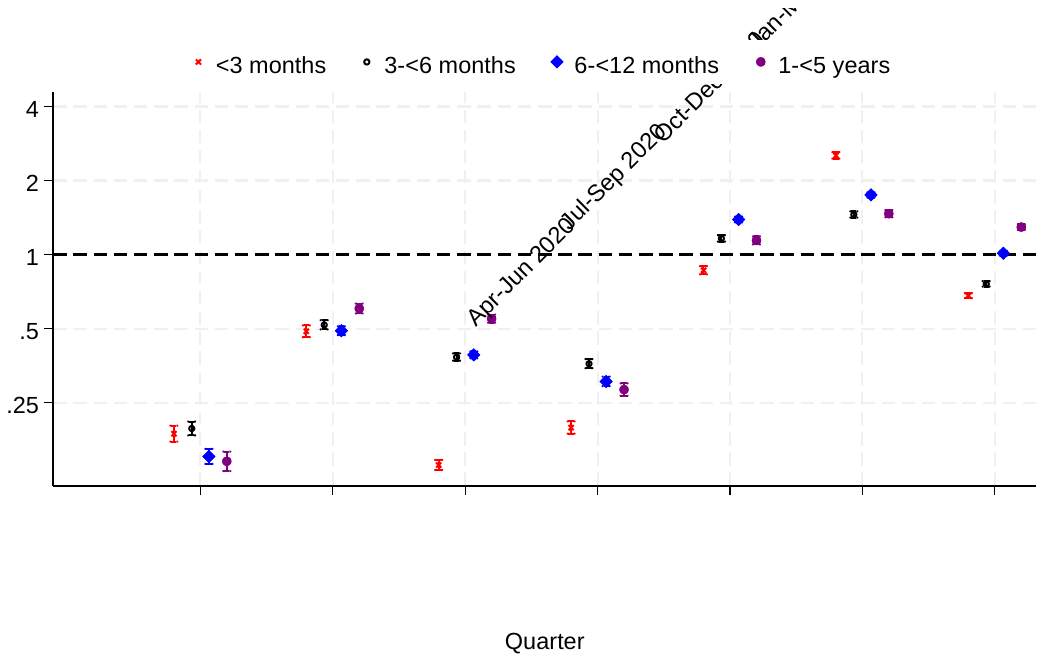 |
| --- | --- |
| 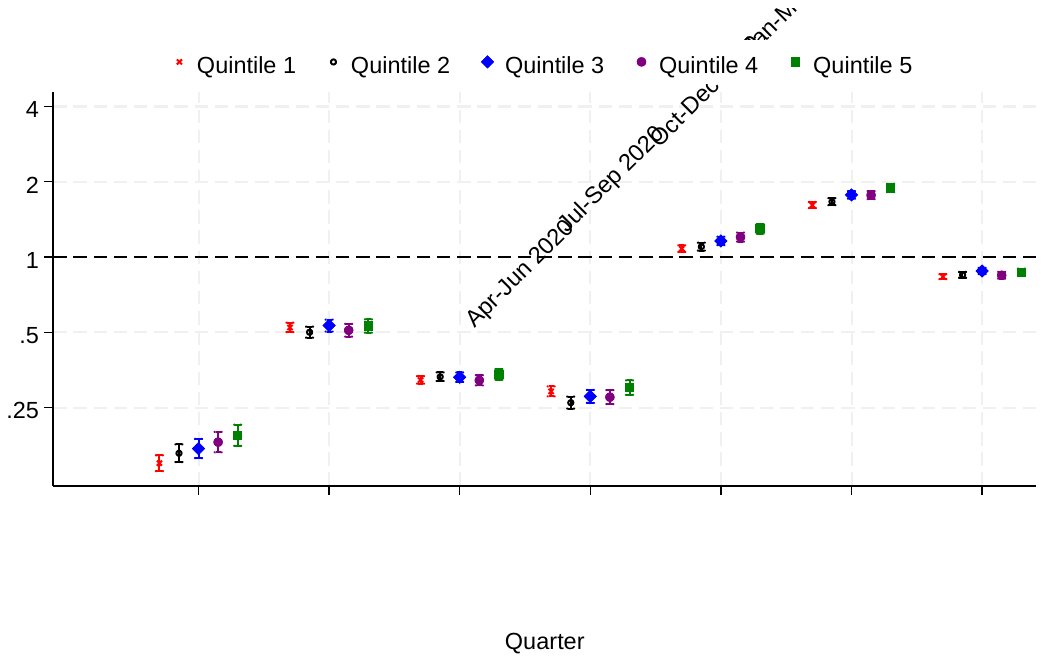 | 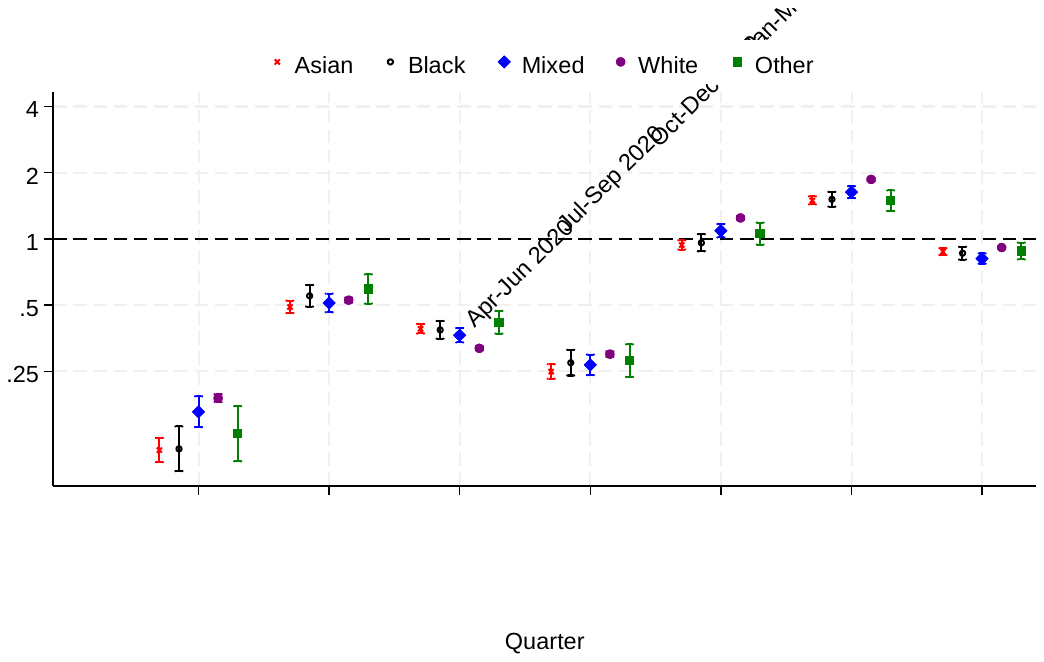 |

*Appendix Figure 24: Incidence rate ratios comparing quarterly respiratory infection admission rates to corresponding pre-pandemic quarters, by preterm birth and presence of congenital anomalies.*

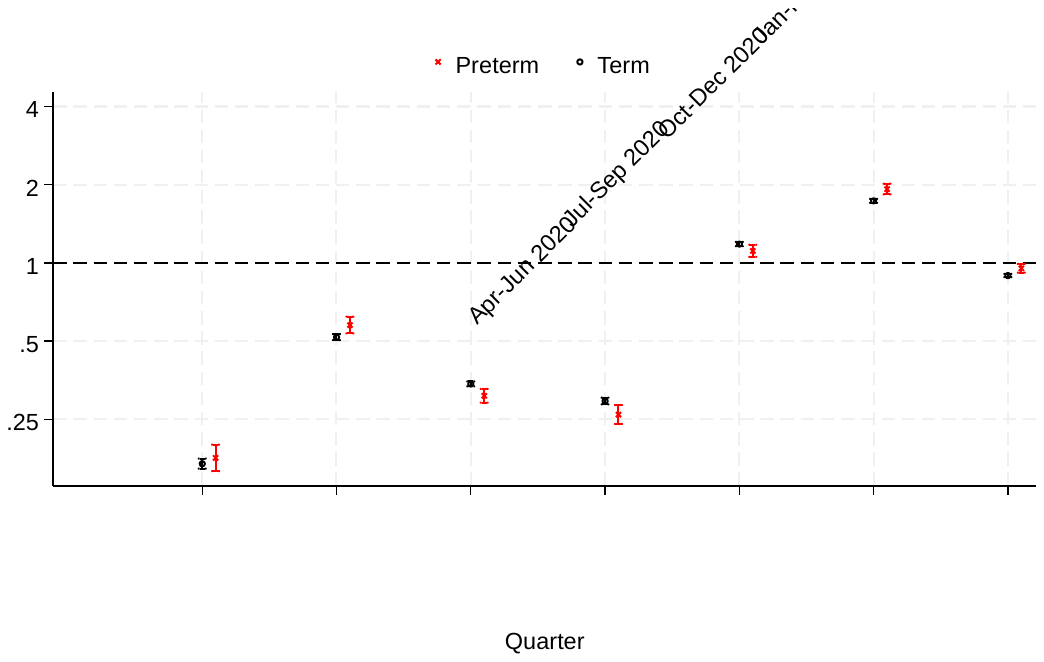

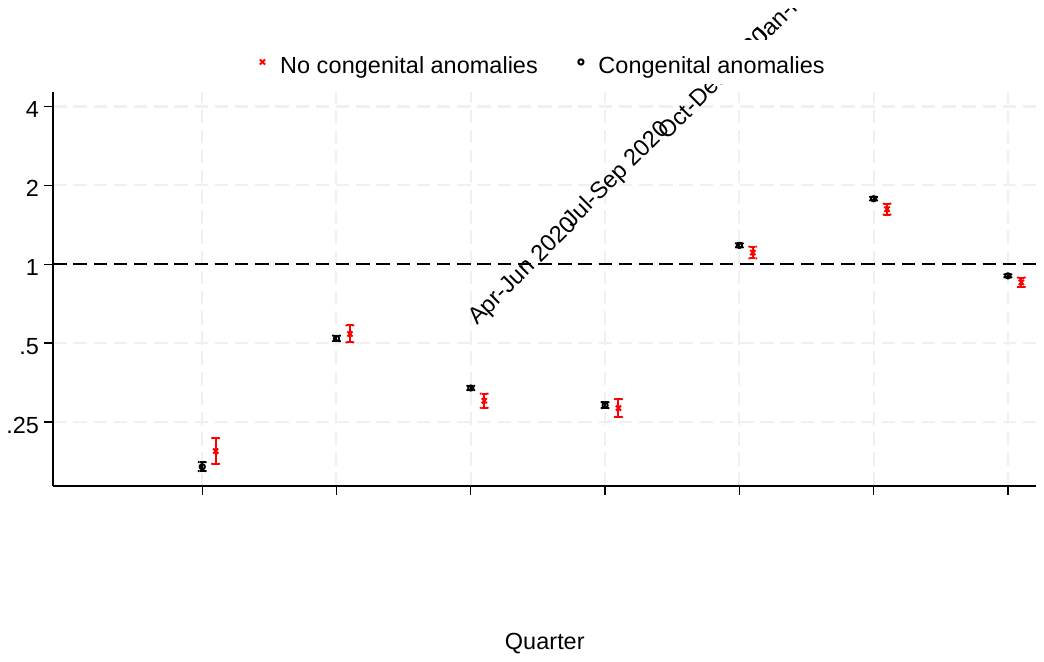

*Appendix Figure 25: Incidence rate ratios comparing quarterly gastrointestinal infection admission rates to corresponding pre-pandemic quarters, by sex, age, area-level deprivation quintile and ethnicity.*

| 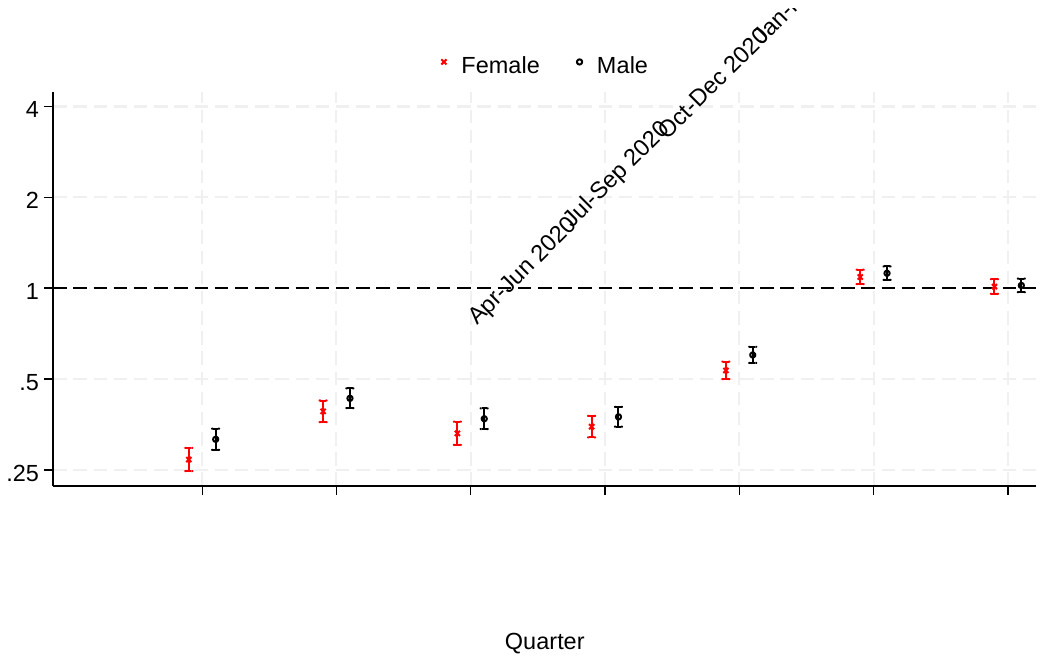 | 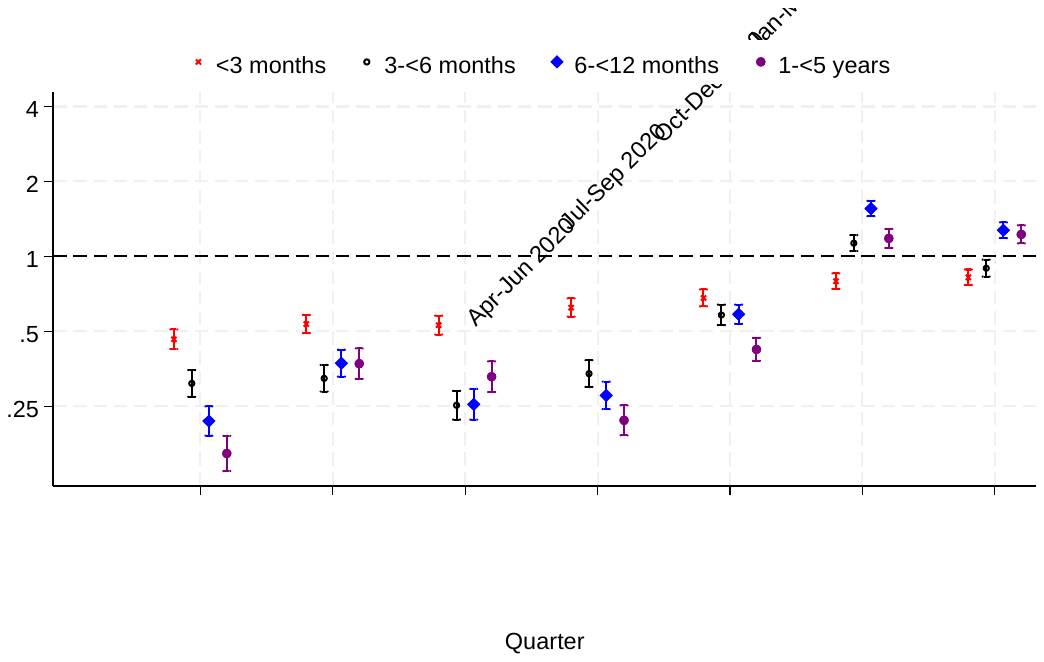 |
| --- | --- |
| 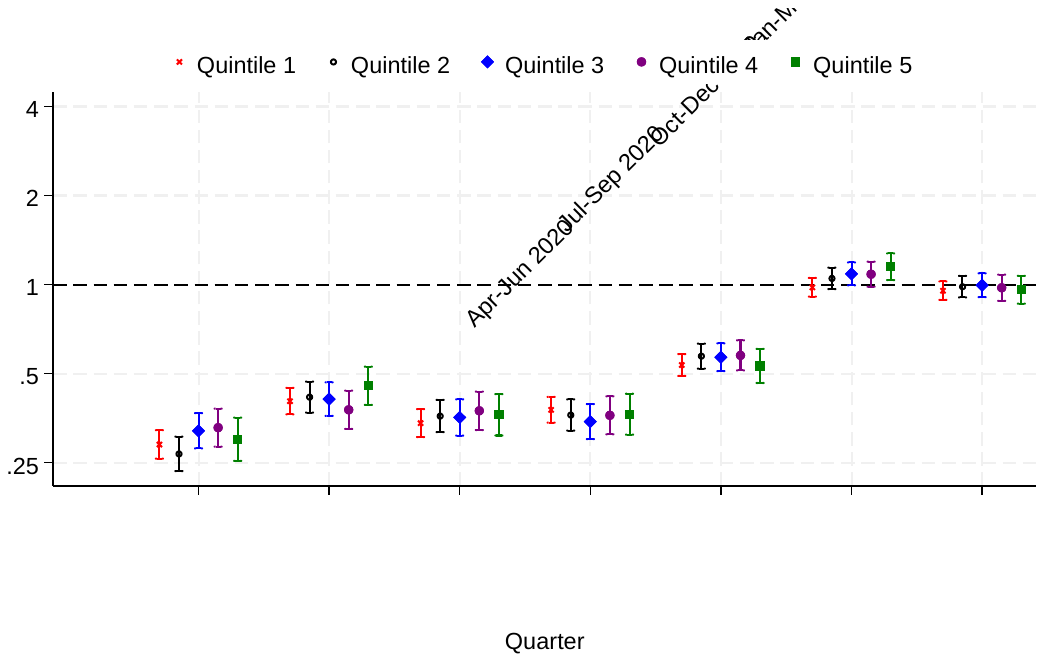 | 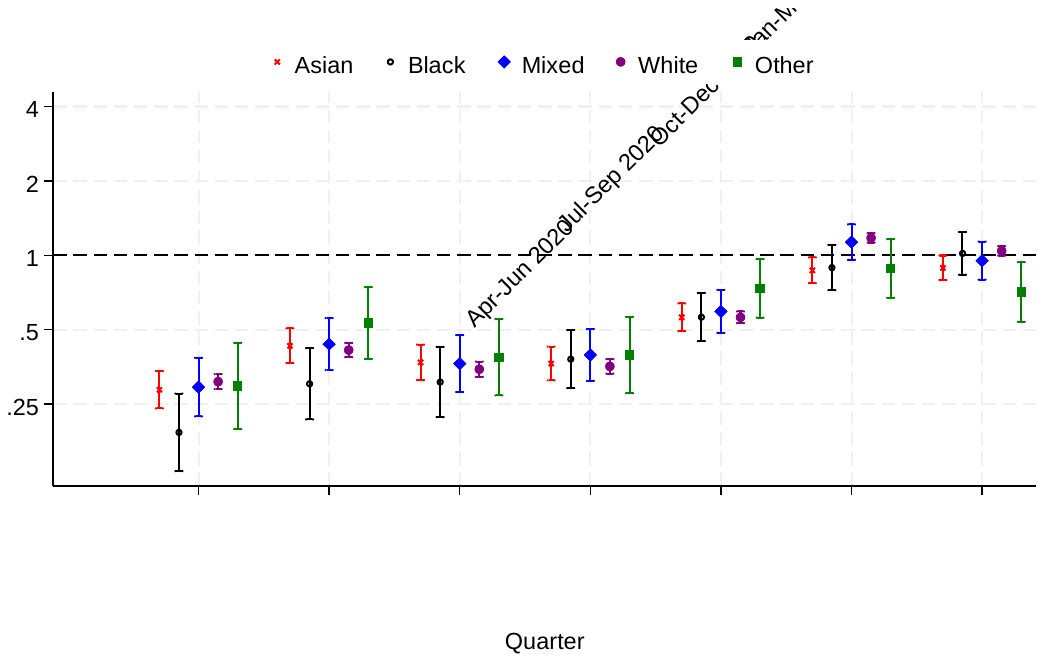 |

*Appendix Figure 26: Incidence rate ratios comparing quarterly gastrointestinal infection admission rates to corresponding pre-pandemic quarters, by preterm birth and presence of congenital anomalies.*

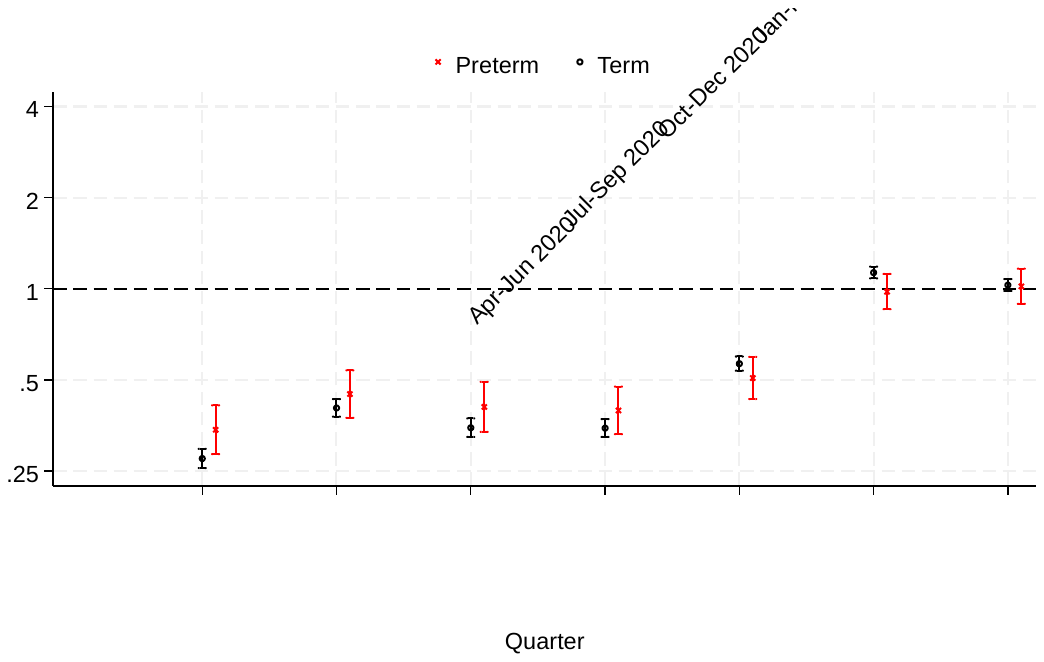

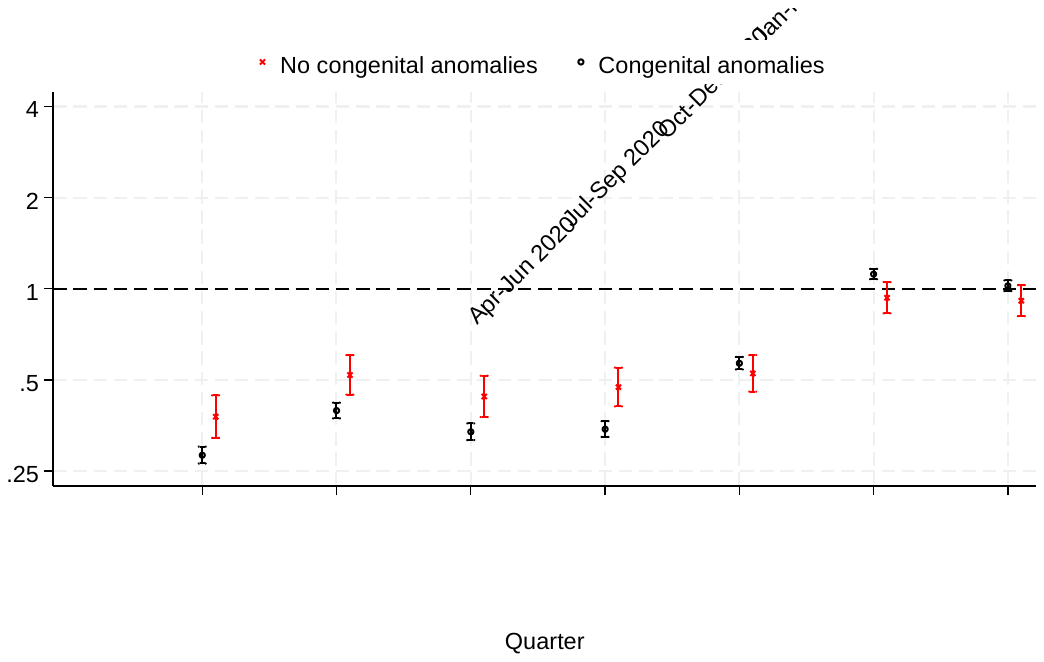

*References*

1.	Kuan, V., et al., *A chronological map of 308 physical and mental health conditions from 4 million individuals in the English National Health Service.* Lancet Digit Health, 2019. **1**(2): p. e63-e77.
